# cfGWAS reveal genetic basis of cell-free DNA features

**DOI:** 10.1101/2024.08.28.24312755

**Authors:** Huanhuan Zhu, Yan Zhang, Shuang Zeng, Linxuan Li, Rijing Ou, Xinyi Zhang, Yu Lin, Ying Lin, Chuang Xu, Lin Wang, Guodan Zeng, Jingyu Zeng, Lingguo Li, Yongjian Jia, Yu Wang, Fei Luo, Meng Yang, Yuxuan Hu, Xiameizi Li, Han Xiao, Xun Xu, Jian Wang, Aifen Zhou, Haiqiang Zhang, Xin Jin

**Author notes:** These authors contributed equally (Huanhuan Zhu, Yan Zhang, Shuang Zeng, Linxuan Li, Rijing Ou). These authors are co-corresponding authors. Correspondence: Xin Jin, Haiqiang Zhang, Aifen Zhou.

## Abstract

cfDNA consists of degraded DNA fragments released into body fluids. Its genetic and pathological information makes it useful for prenatal testing and early tumor detection. However, the mechanisms behind cfDNA biology are largely unknown. In this study, for the first time, we conducted a GWAS study to explore the genetic basis of cfDNA features, termed cfGWAS, in 28,016 pregnant women. We identified 84 significant loci, including well-known cfDNA-related genes DFFB and DNASE1L3, and numerous novel genes potentially involved in cfDNA biology, including PANX1 and DNASE1L1. The findings were further verified through independent GWAS and experimental validation in knockout mice and cell lines. Subsequent analyses revealed strong causal relationships of hematological indicators on cfDNA features. In summary, we presented the first cfGWAS, revealing the genetic basis of cfDNA biology from genome-wide scale. Novel knowledge uncovered by this study keep the promise to revolutionize liquid biopsy technology and potential new drug targeted for certain disease. Given exist of the millions cfDNA whole-genome-sequencing data generated from clinical testing, the potential of this paradigm is enormous.

## Introduction

Cell-free DNA (cfDNA) are DNA fragments released from cells, often originating from processes such as apoptosis, necrosis, and active secretion [1]. These DNA fragments circulate freely in bodily fluids such as blood, saliva, and urine, reflecting the biological status of cells or tissues. CfDNA has extensive applications in non-invasive prenatal testing (NIPT) for detecting fetal abnormalities [2]. It has also emerged as a valuable biomarker for early cancer screening and monitoring [3, 4], as well as for detecting organ rejection in transplantation [5]. Abnormalities in cfDNA has been observed in autoimmune diseases such as systemic lupus erythematosus (SLE) [6]. Additionally, cfDNA shows promise in diagnosing and managing infectious diseases [7, 8].

Commonly used characteristics of cfDNA include cfDNA concentration, fragment length, end motifs, jagged ends, as well as nucleosome footprints [9]. In healthy individuals, cfDNA concentration is low, but it significantly increases in diseases like cancer, inflammation, and tissue damage [10–13]. Therefore, measuring cfDNA concentration can be utilized for disease prediction and monitoring. The fragment length of plasma cfDNA fragments typically ranges from 50 to 600 base pairs; but this may vary due to individual differences, physiological states, and disease conditions [9]. Researchers analyzed the fragment lengths to capture the disease signals, such as aberrant shortening in cfDNA from cancer patients [14, 15]. End motifs refer to the nucleotide composition at the 5’ end of cfDNA, generated during cfDNA digestion. Analyzing these motifs helps trace cfDNA origin and provides insights into physiological and pathological states [16].

These cell-free DNA features arise from the degradation of DNA both inside and outside of cells, and some may correlate due to shared biological processes [9, 17, 18]. Identifying the genes involved in cfDNA fragmentation and their roles is crucial for understanding the mechanisms behind physiological and pathological changes in cfDNA fragmentomic characteristics. However, previous studies have been limited to targeted analysis strategy on limited number of genes [19–22]. To date, only three nucleases, *DFFB*, *DNASE1*, and *DNASE1L3* have been proven to affect cfDNA digestion and fragment characteristics [23]. Studies on nuclease-deficient mice have revealed their role in cfDNA degradation and shaping fragmentomics [10, 24, 25]. It has been demonstrated that cfDNA is first generated intracellularly with *DFFB*, *DNASE1L3*, and other nucleases, followed by extracellular fragmentation with circulating *DNASE1L3* and *DNASE1* [26]. However, the genome-wide genetic basis and other key genes involved in regulating the formation of cell-free DNA are still unknown.

Genome-wide association study (GWAS) is a research method used to identify genetic locus that are statistically associated with specific traits or diseases [27]. Over the past two decades, GWAS have successfully identified hundreds of thousands of variants linked to numerous human traits and diseases, including height, weight, cardiovascular diseases, tumors, and autoimmune diseases [28–30]. As the most comprehensive repository of genotype-trait associations, by May 2024, the GWAS catalog has contained nearly 7,000 publications and over 600,000 association signals, serving as a valuable resource for exploring the genetic background of phenotypes [31].

In previous works, our group has proven that with appropriate algorithms and methodology, cfDNA could be utilized as a resource for high-quality genetic analysis, including variant detection, allele frequency estimation, genetic structure analysis and GWAS [32–36]. In this study, we leveraged genetic and phenotypic data from 28,016 Chinese pregnant women to conduct the first cell-free DNA based genome-wide association study, termed cfGWAS. We used the same copy of whole-genome sequencing data from cfDNA to extract both genotype information and cell-free DNA features, identifying genetic loci associated with these cfDNA molecular characteristics. A comprehensive overview figure has been provided to illustrate the overall design of this work (Figure 1).

**Figure 1.**
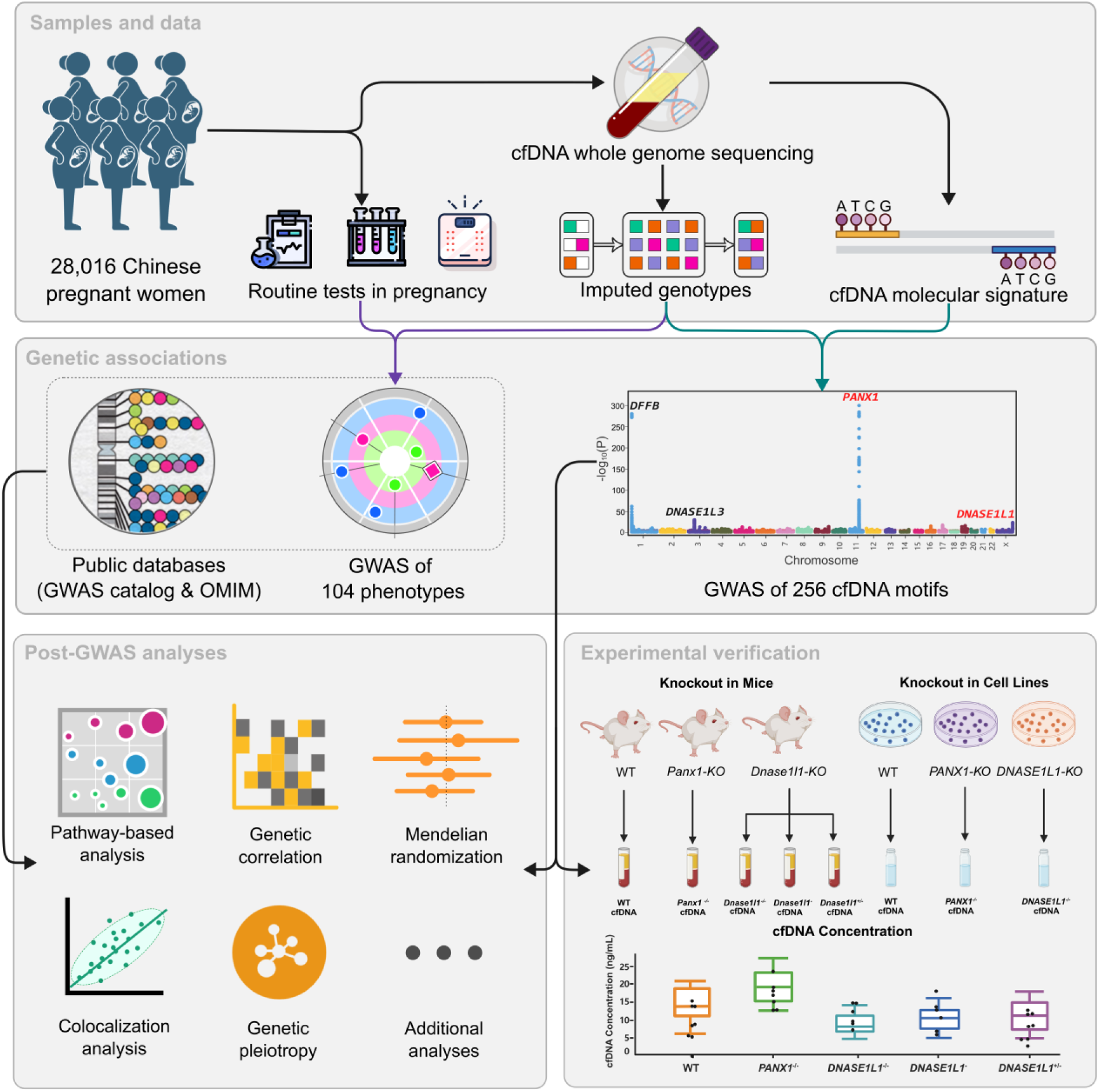
Overview of the study design. A total of 28,016 pregnant women were included in this study, with clinical phenotypes from prenatal care and sequencing data from non-invasive prenatal testing (NIPT) collected. Fragmentation characteristics of cell-free DNA (cfDNA) were extracted from the sequencing data and, after genotype imputation, genotype data were obtained for genome-wide association studies (GWAS). A total of 104 phenotypes and 256 motifs were subjected to GWAS as phenotypic variables, discovering the novel associations of *PANX1* and *DNASE1L1* genes with the cfDNA end motifs. Experiments with gene knockout (KO) mice and cell lines were conducted to validate these associations. Additionally, a series of post-GWAS analyses were performed by integrating the one-sample 104 phenotypes GWAS results, reported associated loci from the GWAS catalog, and diseases reported in the OMIM database, to explore the biological mechanisms, causal relationship and gene pleiotropy of cfDNA fragmentation characteristics.

Key findings from our study include the identification of 84 genome-wide significant loci linked to 218 cell-free DNA end motifs, majority of them were novel associations to cfDNA features, with rediscovery of well-known cfDNA-related genes such as *DFFB* and *DNASE1L3*, and the discovery of novel genes like *PANX1* and *DNASE1L1*. These results were validated through independent replication studies and experimental validation in knockout mice and cell lines. Furthermore, our one-sample integrative analysis revealed strong genetic correlations between cfDNA features and phenotypes such as BMI, weight, white blood cell count, and neutrophil count. Given the widespread clinical use of cfDNA whole-genome sequencing, our work underscores the paradigm-shifting potential of cfGWAS in medical research and practice.

## Results

### Genome-wide association study

#### GWAS results summary

Following the protocol we previously developed, we detailed the genotype imputation performance [34]. To summarize, we utilized NIPT data from 38,668 samples for genotype imputation, achieving an imputation accuracy with an R^2^ value close to 0.8. Covariates included the pregnant woman’s age, gestational week during NIPT screening, sequencing depth, and the first 10 principal components (PCs) of the genotype data. After adjusting for these covariates, the effective sample size was reduced to 28,016, subsequently utilized in the GWAS analysis of cfDNA molecular signatures, focusing on the 4-mer end motifs. At the genome-wide significance threshold of 5e-08, we identified a total of 672 significant locus-motif associations, involving 84 unique loci and 218 end motifs (Table S1). While at the stricter study-wide significance threshold of 1.95e-10 (=5e-8/256), there were 382 significant associations, involving 23 loci and 180 motifs. Note that, all associations that passed the genome-wide significance threshold were maintained for subsequent analyses to ensure the inclusion of all potentially meaningful findings. To assess the reliability of our GWAS results, we drew the Quantile-Quantile plots (QQ-plots) and calculated the genomic inflation factor (GIF) of each motif (Figure S1). Specifically, the GIF values were consistently around 1 across all 256 motifs, indicating no substantial inflations and suggesting well-conducted GWAS analyses.

#### GWAS results visualization

To improve the informative visualization of our GWAS results, we synthesized the summary statistics for the 256 motifs by selecting the minimum p-value for each SNP across all motifs, thereby generating an integrated GWAS summary dataset. This integrated GWAS summary data was then used to generate a Manhattan plot, providing a comprehensive overview of the results (Figure 2a). The most significant locus identified is *PANX1*, with the lead SNP rs76201528 (p-value = 3.93e-350). Following *PANX1* is *DFFB*, with lead SNP rs74752626 (p-value = 5.14e-281). The subsequent two most significant loci were *DNASE1L3* (rs12633655, p-value = 4.82e-31) and *DNASE1L1* (rs2283762, p-value = 1.47e-24). For more details of each signal, we provided the regional plots (Figure S2). Other significant loci include *PSMD3*, *FXYD5*, *PADI4*, *ABO*, *FCHO2*, *MSR1*, and so on. For each locus, we counted the number of its associated motifs and presented a list of loci with more than five associated motifs (Figure 2b). The top five loci with the largest number of associated motifs were *PANX1* (145), *DFFB* (143), *ANKRD26P1* (39), *DNASE1L3* (34), and *PSMD3* (29). From the perspective of end motifs, we summed up the number of significant loci for motifs with the same starting nucleotide. The counts for ‘A’, ‘T’, ‘C’, and ‘G’ were 175, 143, 213, and 141, respectively. Furthermore, we used a Venn diagram to visualize the shared and unique significant loci associated with four motif groups (A, T, C, and G) (Figure 2c). This analysis revealed seven loci common to all motif groups, including *PANX1*, *DFFB*, *DNASE1L3*, and *PADI4*. The numbers of exclusively associated loci for ‘A’, ‘T’, ‘C’, and ‘G’ were 16, 3, 31, and 8, respectively.

**Figure 2.**
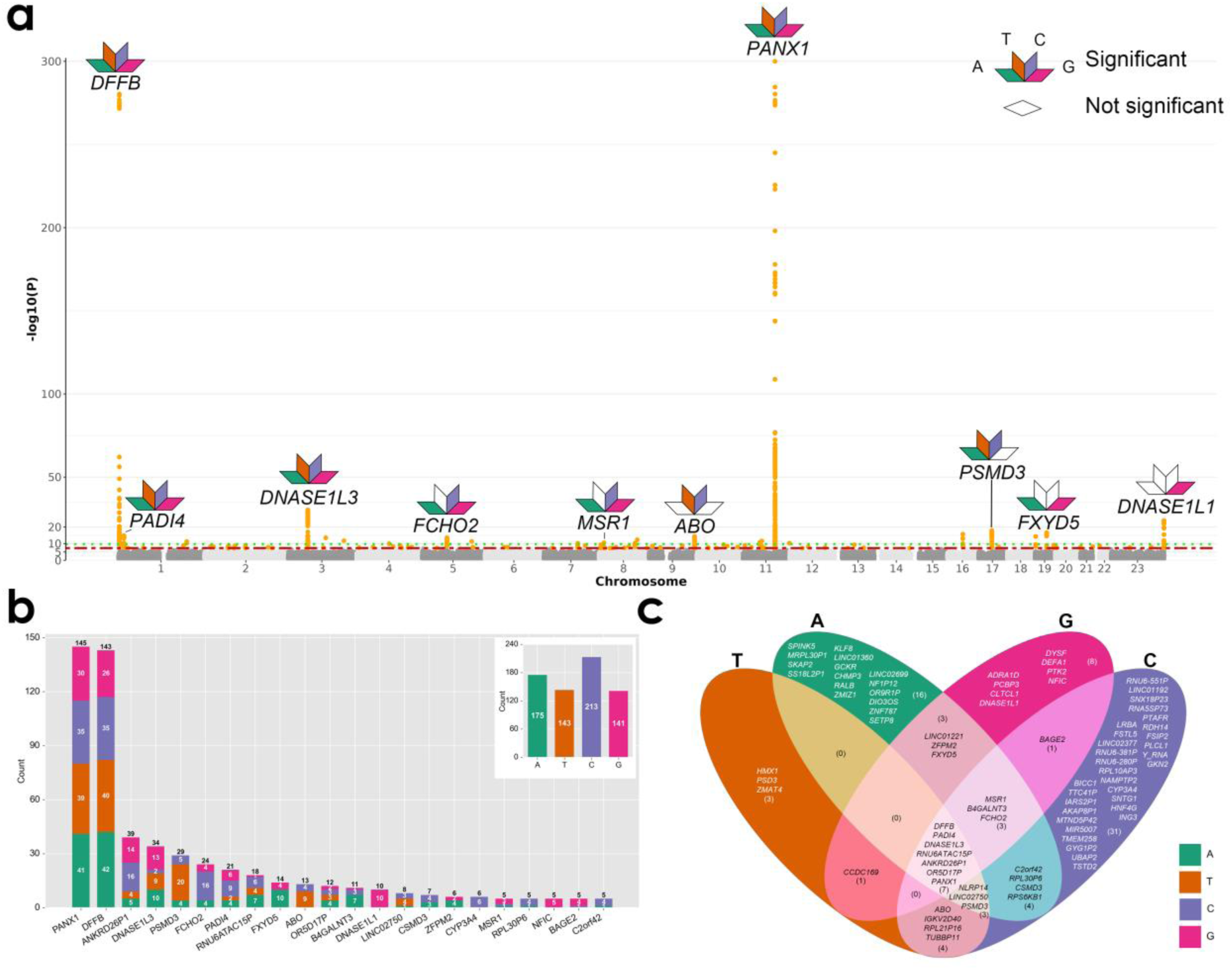
The results for GWAS analysis of 256 motifs. a) a comprehensive Manhattan plot integrating the GWAS results for 256 motifs, each dot represents the minimum p-value of a SNP associated with the 256 motifs; several gene names are annotated at the top of significant loci; the association of motifs beginning with the four nucleotide bases A, T, C, and G with these loci indicated by differently colored diamonds, b) a bar plot presents the number of associated motifs that hit on each locus, with the frequency of each start nucleotide A, T, C and G differed by distinct color, c) the Venn diagram depicting the unique and shared loci hit by motifs starting with each nucleotide base (A, T, C and G).

#### Biological implications

To explore the biological implications of our findings, we examine the functions of four prominent loci: *PANX1*, *DFFB*, *DNASE1L3*, and *DNASE1L1*. The gene *PANX1* encodes Pannexin-1, a protein that forms channels within the cell membrane, facilitating the exchange of molecules between the intracellular and extracellular environments [37]. These channels play diverse roles in various cellular processes, encompassing cell signaling, apoptosis, inflammation, and development [38]. On the other hand, the gene *DFFB*, also referred to as DNA fragmentation factor subunit beta, encodes a protein integral to apoptosis, or programmed cell death. Specifically, *DFFB* is a pivotal component of the DNA fragmentation complex, crucial for cleaving DNA into fragments during apoptosis [26].

Moving on to the gene *DNASE1L3*, it encodes deoxyribonuclease 1-like 3, belonging to the deoxyribonuclease I family. *DNASE1L3* is predominantly expressed in the spleen and lymph nodes and plays a pivotal role in DNA degradation from apoptotic cells, thereby aiding in the clearance of cellular debris [26]. This process is paramount for maintaining tissue homeostasis and averting autoimmune responses triggered by the release of self-DNA. Similarly, the gene *DNASE1L1* encodes deoxyribonuclease 1-like 1, another member of the deoxyribonuclease I family. *DNASE1L1* is instrumental in the degradation of DNA from apoptotic cells and extracellular DNA, facilitating the clearance of cellular debris and maintaining tissue homeostasis [39].

Both *DFFB* and *DNASE1L3* are well-established apoptotic nucleases, highlighting their significant roles in cellular homeostasis, apoptosis and cfDNA fragmentation [26]. As novel discoveries, our primary attention shifts towards investigating the genes *PANX1* and *DNASE1L1* for further analysis. To verify our findings of the potential genetic effects of these four genes on cfDNA molecular signatures, we conducted a GWAS replication study utilizing an independent cohort of natural populations and also performed experimental verifications using both knockout mice and knockout cell lines.

### Replication study with independent samples

To validate our GWAS findings of cfDNA end motifs, we conducted a replication analysis using an independent cohort of 442 subjects. This analysis successfully replicated two significant signals: *DFFB* and *PANX1* (Figure S3). For the *DFFB* locus, the lead SNP, rs35265408, exhibited a p-value of 7.07e-13 in the replication study. We note that this SNP was removed due to Hardy– Weinberg equilibrium filtration in our discovery study. Furthermore, the lead SNP identified in the discovery study for the *DFFB* locus was also found to be significant in the replication study (p-value = 7.37e-10). In the *PANX1* locus, the lead SNP identified was rs1946143444 (p-value = 6.31e-10). However, this SNP is an insertion-deletion (INDEL) and was not analyzed in the discovery study. The successful replication of the *DFFB* and *PANX1* loci in an independent cohort provides strong evidence for their association with cfDNA molecular signatures. Although the small sample size limited our ability to fully replicate the findings for *DNASE1L3* and *DNASE1L1*, this study contributes valuable insights and further strengthens the reliability of our cfGWAS results.

### Experimental verifications

#### CfDNA concentration in *PANX1* knockout mice and cell lines

To validate the role of the newly identified genes in the generation and clearance of cell-free DNA, we homozygously knocked out (KO) the *Panx1* gene in mice. The plasma cfDNA concentrations in *Panx1* KO mice (n=6) were compared with those of wild-type (WT) mice (n=6) (STAR Methods). The fold change of cfDNA concentration in each mouse relative to the average cfDNA concentration in WT mice was calculated. As depicted in Figure 3, we observed a significant increase in the fold change of cfDNA concentration in *Panx1* KO mice (median: 1.63, range: 1.30-2.90) compared to WT mice (median: 0.94, range: 0.69-1.43, p-value=0.019), suggesting the knockout of *Panx1* gene influenced the release of cfDNA into plasma (Figure 3a).

**Figure 3.**
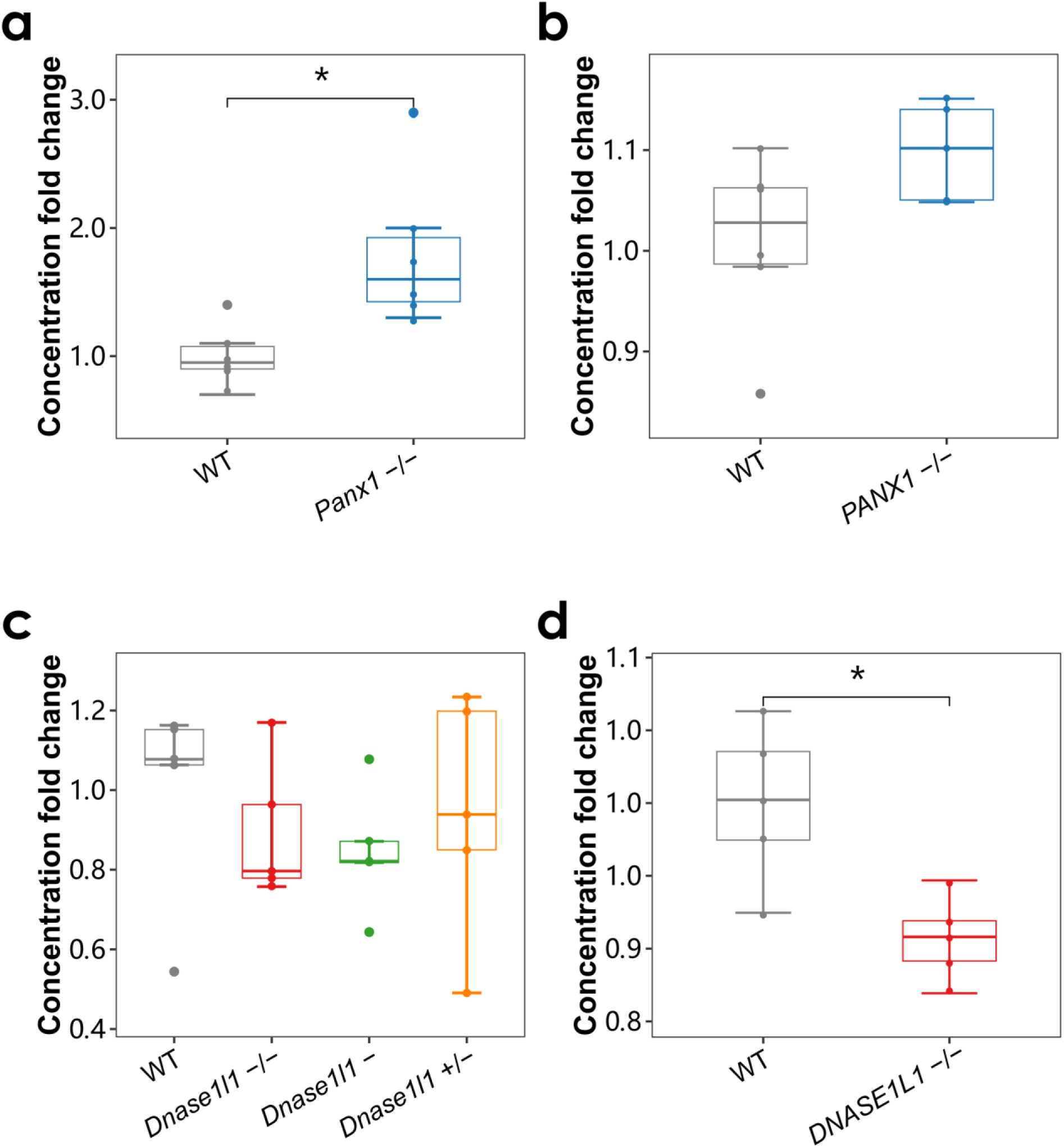
The results for experimental validation. The box plots presenting the distribution of cfDNA concentration fold changes for a) plasma samples from wild-type (WT) and *Panx1* KO mice; b) cell culture supernatant from WT and *PANX1* KO cell lines; c) plasma samples from wild-type (WT) and *Dnase1l1* KO mice; d) cell culture supernatant from WT and *Dnase1l1* KO cell lines. Since the *Dnase1l1* gene is located on the X chromosome, we generated three types of *Dnase1l1* KO mice. Significant differences with p-value < 0.05 between two groups are marked with asterisks (*).

Furthermore, we knocked out the *PANX1* gene in Jurkat Cell line. The cfDNA concentration in the cell culture supernatant of the *PANX1* KO cells (5 replicates) was compared to that of WT Jurkat cells (5 replicates). Although the difference did not reach statistical significance, we do observe a trend of increased cfDNA concentration of *PANX1* KO cells (median fold change: 1.10, range: 1.05-1.15) compared to the WT cells (median fold change: 0.99, range: 0.86-1.10, p-value=0.08, Wilcoxon test) (Figure 3b). The results of both *in vivo* and *in vitro* KO experiments suggest that the *PANX1* gene plays a crucial role in the generation of cfDNA, and the knockout of the *PANX1* gene leads to an increased release of cfDNA.

#### CfDNA concentration in *DNASE1L1* knockout mice and cell lines

Similarly, we conducted knockout of the *Dnase1l1* gene in mice and quantified the plasma cfDNA concentration in both WT and KO mice. Since the *Dnase1l1* gene is located on the X chromosome, we generated three types of *Dnase1l1* KO mice: homozygous KO mice (*Dnase1l1* −/−, n=5), heterozygous KO mice (*Dnase1l1*-/+ n=5), and hemizygote KO mice (*Dnase1l1* –, n=5). The plasma cfDNA concentration in these KO mice was compared to that in WT mice (n=5). As shown in Figure 3c, a consistent decrease in the fold change of cfDNA concentration was observed in *Dnase1l1* −/− mice (median: 0.80, range: 0.76-1.17, p-value=0.47), *Dnase1l1* –/+ mice (median: 0.94, range: 0.49-1.23, p-value=0.75), and *Dnase1l1*-mice (median: 0.82, range: 0.64-1.08, p-value=0.29) compared to WT mice (median: 1.08, range: 0.54-1.16). Furthermore, we homozygously knocked out the *DNASE1L1* gene in HEK293 cells and compared the cfDNA concentration in the supernatant with that of wild-type HEK293 cells (5 replicates). Significantly decreased fold change of cfDNA concentration was observed in *DNASE1L1* KO cells (median: 0.91, range: 0.87-0.95) compared to WT cells (median: 1.00, range: 0.92-1.06, p-value=0.014) (Figure 3d). The results obtained from both *in vivo* and *in vitro* knockout experiments provide compelling evidence supporting the crucial role of the *DNASE1L1* gene in the generation of cfDNA, and the knockout of the *DNASE1L1* gene leads to a decreased release of cfDNA.

### Post-GWAS analyses of motif summary statistics

#### Heritability

Heritability measures the proportion of phenotypic variance explained by genotypes, thus quantifying the genetic influence on phenotypes. We calculated the heritability of each motif using LD score regression (Table S2, Figure S4). Notably, among the 256 motifs, 14 motifs exhibited a heritability exceeding 10%, while 69 motifs fell within the range of 5% to 10%, suggesting a substantial genetic component in their variation. We also provide the number of significant loci associated with each motif (Table S2, Figure S4). As expected, the heritability is generally positively proportional to the number of associated loci.

#### Heritability partition

Our partitioning heritability analysis revealed significant contributions of cell type-specific elements to the heritability of cfDNA end motifs. At a suggestive significance threshold of 1e-2, we identified 290 significant cell-type-motif associations, involving 87 unique cell types and tissues (Tables S3-S4, Figure S5a). Interestingly, the most frequently implicated cell types in these associations were phagocytes (23 motifs), followed by neutrophils (18 motifs), bone marrow cells (13 motifs), and synovial fluid (10 motifs), all belonging to the blood/immune category. The most prominent association is observed between the GATC motif and neutrophils (p-value=7.32e-5), and others include CTCA motif and phagocytes (p-value= 5.68e-4), and GACC and neutrophils (p-value= 8.65e-4) (Figure S5b). These findings align with the known role of phagocytes (including neutrophils and monocytes) as both sources and clearers of cfDNA during immune responses [40]. Our analysis identified an enrichment of hematological cell types, particularly immune cells, in the heritability of cfDNA motif features, indicating their active involvement in cfDNA processing.

#### Pathway-based association analysis

The pathway-based analysis results of all 256 end motifs were provided in Supplementary Table S5. At a suggestive significance threshold of 1e-3, a total of 732 motif-pathway pairs demonstrated significance, and they were distributed among four motif groups starting with ‘A’, ‘T’, ‘C’, and ‘G’, associating with 190, 311, 148, and 83 pathways, respectively (Table S6, Figure S6a). These significant relationships encompassed 101 unique pathways, categorized into groups such as cell death and apoptosis, immune response, cancer-related pathways, among others.

Among the identified pathways, the top four with the most significant motifs included the Biocarta SET pathway leading with 80 associated motifs, followed by Biocarta DNA fragment pathway (75 motifs), Reactome apoptosis-induced DNA fragmentation (62 motifs), and Biocarta Mitochondria pathway (42 motifs). Specifically, the Biocarta SET pathway is vital in regulating B cells by providing crucial survival signals that prevent inappropriate apoptosis [41]. Both the Biocarta DNA fragment pathway and Reactome apoptosis-induced DNA fragmentation are implicated in DNA breakdown, a pivotal process in apoptosis regulation and cell death [42]. Furthermore, the Biocarta Mitochondria pathway sheds light on how mitochondria contribute to cellular functions like metabolism and apoptosis.

Moreover, we provided the distribution of pathway-based analysis p-values for all motifs through Boxplots, sorted by the median (Figure S6b). Notably, the top pathways are consistent with the above observations, including the Biocarta SET pathway, Biocarta DNA fragment pathway, Biocarta Mitochondria pathway, Biocarta caspase pathway, and Reactome apoptosis-induced DNA fragmentation. In conclusion, the pathway-based association analysis illuminated the biological processes such as DNA fragmentation, apoptosis, and Mitochondria pathway in governing cfDNA generation and clearance.

### Post-GWAS analyses with one-sample pregnancy phenotypes

#### Genetic correlation

In a parallel study with the same group of pregnant women, we have performed genetic analysis on their 104 pregnancy phenotypes, including laboratory tests during pregnancy (e.g., hematology, liver-function), maternal information (e.g., height, BMI), and neonatal outcomes (e.g., birthweight, birth length) [34]. We conducted a genetic correlation analysis between the 256 end motifs and the 104 pregnancy phenotypes to assess the overall genetic similarity between each pair (Tables S7-S8, Figure 4a). At the suggestive significance threshold of 1e-3, a total of 61 motif-phenotype pairs exhibit significant correlations (Table S9, Figure S7). These pairs involve 37 unique cfDNA end motifs and 9 pregnancy phenotypes. The A-, T-, C-, and G-end motifs have 9, 27, 15, and 10 significant pairs, respectively. The 9 phenotypes are maternal BMI (19 motifs), maternal weight (14 motifs), white blood cell count (7 motifs), neutrophils count (6 motifs), uric acid (6 motifs), neutrophils percentage (5 motifs), lymphocytes percentage (2 motifs), aspartate transferase levels (1 motif), and platelet count (1 motif).

**Figure 4.**
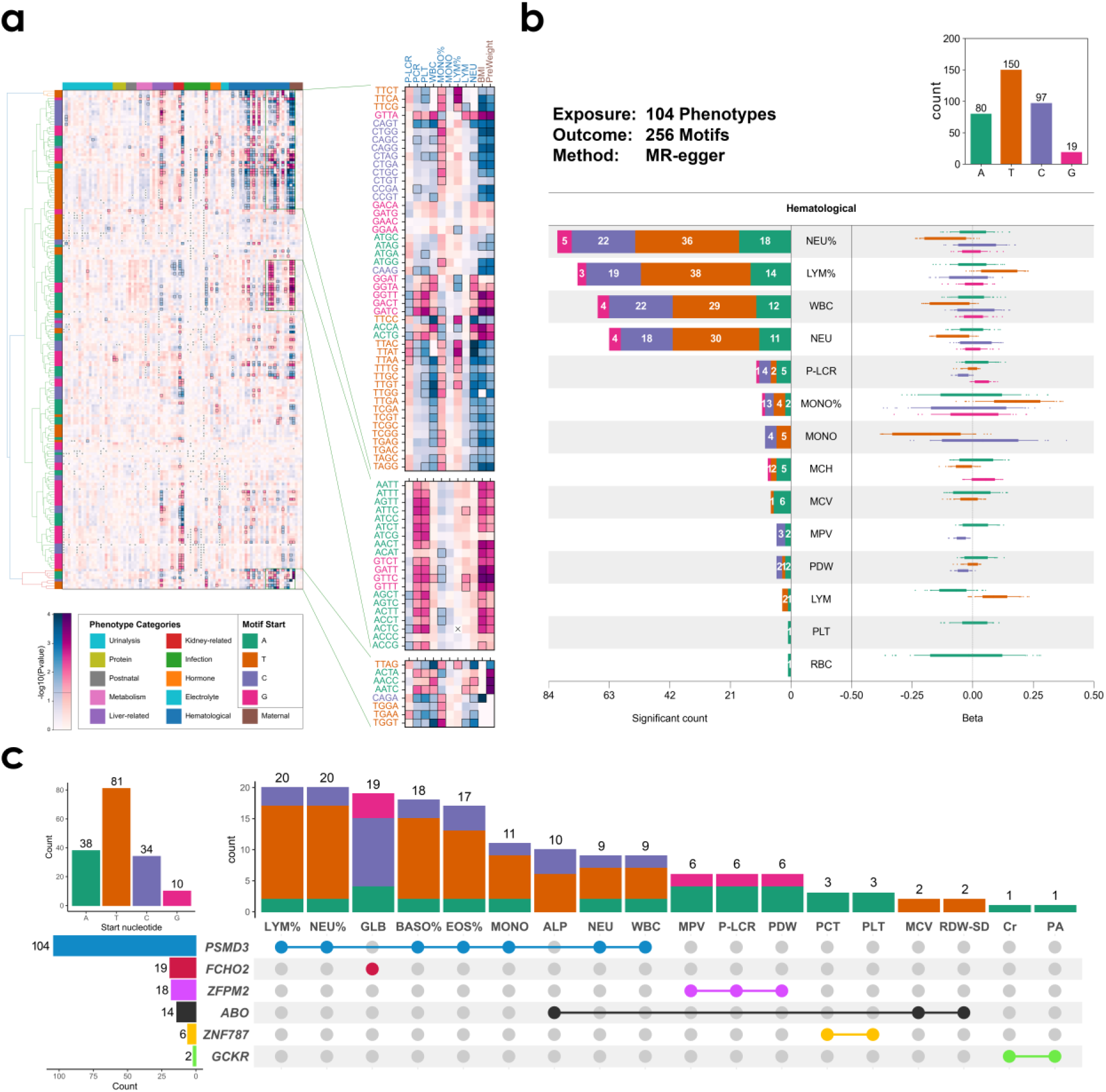
The results for one-sample post-GWAS analyses. a) A heatmap illustrating the pairwise genetic correlation p-values between 104 pregnancy phenotypes and 256 motifs, positive and negative genetic correlations are represented in red and blue, respectively. The motifs have been hierarchically clustered, and pregnancy phenotypes are sorted by category; different phenotype categories are distinguished by color coding on the axes; Several regions with strong genetic correlations are magnified and displayed on the right, b) The results for Mendelian randomization analysis, highlighting the 14 hematological phenotypes that exhibit significant causal relationships with motifs; motifs are categorized into four groups (A, T, C and G) according to the start nucleotide; for each motif group, the beta values from the MR-Egger analysis are depicted in boxplots on the right side, while the number of significant motifs is illustrated in bar charts on the left side, and the total number of significant motifs across each group is displayed in bar charts at the top right corner, c) the results for colocalization analysis between motifs and pregnancy phenotypes, for each significance locus, if the locus is also present in the GWAS results of pregnancy phenotypes, it is denoted by a colored large dot marker; the bar plots is used to display the number of co-localized motifs in each phenotype above the dot matrix, with different colors distinguishing motifs from different groups (A, T, C and G); on the right side of the dot matrix, the total number of motifs co-localized with phenotypes for each locus is shown; on the upper left corner of the dot matrix, the total number of co-localized motifs across each motifs group is displayed.

The relationship between BMI, weight, and cfDNA during pregnancy has been extensively researched. Multiple studies have identified an association between increasing maternal weight and a decrease in cell-free fetal DNA levels [43–46]. Additionally, other studies have shown that total cfDNA levels are elevated in obese pregnant women, primarily due to increased necrosis and apoptosis of adipose tissue [47–49]. Uric acid (UA), a byproduct of purine metabolism, is generated from the breakdown of nucleic acids (DNA and RNA) and ATP [50]. Elevated UA levels are closely linked to oxidative stress, playing a significant role in renal tubular epithelial cell injury and apoptosis [51, 52]. A recent study also suggests that uric acid may induce mitochondrial dysfunction and apoptosis through the downregulation of mitochondrial phosphatidylserine decarboxylase [53]. We will discuss the relationship between hematological indicators and cfDNA in the next section.

#### Mendelian randomization

We conducted bi-directional Mendelian randomization analyses to explore the potentially causal relationships between pregnancy phenotypes and end motifs. Initially, we treated pregnancy phenotypes as exposures and motifs as outcomes (denoted as P->M) and subsequently reversed the roles (denoted as M->P). At the suggestive significance threshold of 1e-3 based on both MR-Egger method and inverse variance weighted method, our P->M analysis identified 346 potentially causal associations involving 14 pregnancy phenotypes, all belonging to the hematological category (Tables S10-11, Figure 4b). In detail, 307 out of 346 (88.7%) of the identified causal relationships was found between leukocytes (e.g., neutrophils, lymphocytes, and monocytes) and end motifs, followed by thrombocytes (23 causalities) including platelet larger cell ratio and mean platelet volume, as well as erythrocytes (16 causalities) including mean corpuscular hemoglobin and mean corpuscular volume.

The release of cfDNA and its relationship with leukocytes has been extensively studied and can be understood through the context of cellular apoptosis, infection and inflammatory responses, and DNA clearance mechanisms [54, 55]. The leukocytes, particularly neutrophils, are the major contributors to cfDNA in cancer samples, accounting for approximately 76% of cfDNA [56]. Specifically, activated neutrophils can undergo a specialized form of cell death called NETosis, a program for formation of neutrophil extracellular traps (NETs), significantly raising cfDNA levels [57]. Moreover, cfDNA released during exercise mainly originates from extramedullary polymorphonuclear neutrophils, influenced by physical impact, low oxygen levels, and elevated core body temperature [58].

Additionally, a recent study discovered that thrombocytes contain genomic DNA fragments from megakaryocytes, and as precursors of thrombocytes, megakaryocytes contribute significantly to cfDNA, accounting for approximately 26% in healthy individuals [59]. Furthermore, the elevated levels of plasma cfDNA have been observed in venous thromboembolism (VTE) patients [60]. The relationship between erythrocytes and cfDNA has been explored. First, the discarded DNA resulting from the maturation process of erythrocytes is a major source of homeostatic cfDNA [61]. Second, a recent study revealed that mature erythrocytes contain long DNA fragments [62]. Consequently, the origin of erythrocytes may also contribute cfDNA to the plasma. Together, these processes shed light on the dynamic regulation of cfDNA by different blood cell types.

For motifs starting with ‘A’, ‘T’, ‘C’, and ‘G’, they were causally affected by 80, 150, 97, and 19 phenotypes, respectively. We additionally provided the fitted linear graphs for causal pairs of phenotypes and motifs based on the MR-Egger method (Figure S8). The lines exhibit intercepts close to zero, suggesting the absence of pleiotropic effects and reliable causal effects in MR analysis.

To ensure the validity of our P->M MR results, we performed reversed M->P analysis, in which the cfDNA motifs as exposures and the phenotypes as outcomes. At the same suggestive significance threshold of 1e-3, we found no causal effects of motifs on phenotypes, with the smallest p-value being 1.32e-3 (ACTT->total protein). This phenomenon is consistent with our prior knowledge that cfDNA features are often considered as consequences rather than causes; the occurrences of some biological or physiological processes (e.g., immune response) may result in alteration of hematological tests (e.g., neutrophils count), causing the change of cfDNA molecular signatures (e.g, concentration, motifs). To further ensure the robustness of the 346 causal associations, we performed genetic pleiotropy and heterogeneity tests (Table S11). At the significance threshold of 1e-3, all the causal relationships are not affected by genetic pleiotropy, with the smallest p-value being 4.81e-3. Even though five associations show potential heterogeneity effects (p-value<1e-3), these effects appear to be driven by a small subset of the genetic variants and do not substantially impact the overall findings.

Our bi-directional MR analysis, which controlled for both genetic pleiotropy and heterogeneity effects, highlighted the causal effects of hematological indicators, especially immune cells, on cfDNA molecular features.

#### Colocalization analysis

To identify shared causal genetic effects on both end motifs and pregnancy phenotypes, we conducted GWAS-GWAS colocalization analysis. This analysis revealed 163 colocalized motif-phenotype pairs at a posterior probability of H4 (one common causal variant) greater than 0.75, distributed across six loci: *PSMD3* (104 pairs), *FCHO2* (19 pairs), *ZFPM2* (18 pairs), *ABO* (14 pairs), *ZNF787* (6 pairs), and *GCKR* (2 pairs) (Table S12, Figure 4c, Figure S9). Notably, the phenotypes with the highest number of colocalized signals were primarily related to hematological traits (leukocytes, thrombocytes, and erythrocytes) and protein levels (globulin and prealbumin).

The most frequent colocalizations appear in locus *PSMD3* linking cfDNA motifs and leukocytes cells. The gene *PSMD3* (proteasome 26S subunit, ATPase, 3) plays critical roles in process of protein degradation, in maintaining cellular health, and in regulating various cellular processes. Silencing of *PSMD3* inhibits cell proliferation and induces apoptosis, as evidenced by the significant reduction in breast cancer cell proliferation and colony formation following *Psmd3* knockdown [63]. Previous GWAS studies have extensively reported its associations with leukocytes cells [64–67] and immune diseases (e.g., asthma, allergic disease) [68, 69].

Additionally, from the perspective of end motifs, T-end exhibited the largest number of colocalized signals (81), followed by A-end (38), C-end (34), and G-end (10). These findings provide evidence for shared genetic background influencing both cfDNA end motifs and pregnancy phenotypes. The significant enrichment of hematological and protein-related traits in the colocalized signals suggests their close relationship with cfDNA molecular features.

### Pleiotropy analyses with public databases

#### GWAS catalog

We further compared our results with the GWAS catalog containing all reported genome-wide significant associations with all possible phenotypes to date. For each trait, we defined a ratio as number of associated loci shared with cfGWAS result in this study over total number of associated loci in GWAS catalog (Table S13). We provided a bubble plot to present the pleiotropic results with GWAS catalog (Figure 5a). Overall, traits with large ratios are mainly belong to hematology and lipid/lipoprotein measurements. Specifically, among traits with more than 50 associated loci, uric acid has the largest ratio, followed by platelet counts, cholelithiasis, and basophils counts. Other traits include height, BMI, asthma, allergic rhinitis, and thrombosis. The relationships between most of these traits and cfDNA molecular features have been discussed in earlier sections.

**Figure 5.**
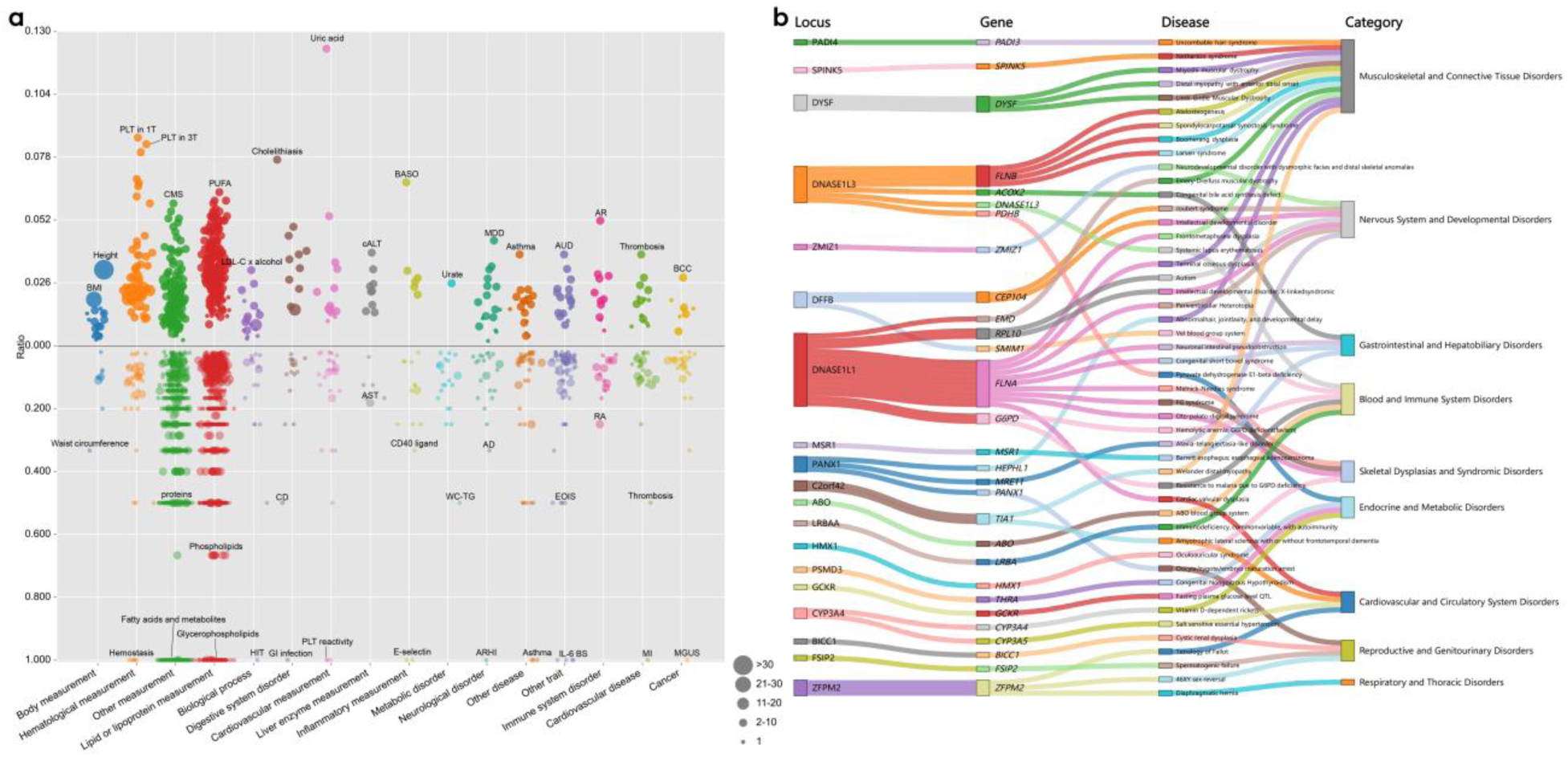
The pleiotropy analysis results with GWAS catalog and OMIM database. a) the bubble plot illustrating the summary of the motif associated loci that overlap with reported association in the GWAS catalog; the top section represents phenotypes with more than 50 variants records in the GWAS catalog, while the bottom section represents those with fewer than 50 records; the size of each bubble corresponds to the number of overlapping variants, and the absolute value of the y-axis coordinate represents the proportion of overlapped variants among all reported variants within each phenotype; the full names of the listed abbreviations of the traits are provided in Table S13 with the abbreviations in square brackets; b) the Sankey diagram, from left to right, sequentially displays the names of significant loci of motifs, the genes contained within each locus, and the corresponding relationships between each gene and the diseases and their categories recorded in the OMIM database.

A large number of traits with less than 50 associated loci have one or two significant loci that all shared with the cfGWAS signals, resulting in the ratio of 1, such as a wide spectrum of glycerophospholipids in the lipid/lipoprotein measurement, hemostasis in hematology, and fatty acids and metabolites in other measurement. Some studies have reported that elevated maternal lipid levels during pregnancy, such as increased triglycerides, may reduce the fetal fraction of cfDNA while increasing the total cfDNA levels [70, 71]. Our findings support aforementioned discoveries that cfDNA molecular features may have close correlation with traits like BMI, hematology, and immune responses from the perspective of genetics.

#### OMIM compendium

Of the 144 mapped genes with significant SNPs associated with cfDNA end motifs, 48 have documented disease associations according to the OMIM database (Table S14, Figure 5b). These associated diseases are distributed across nine organ systems, with musculoskeletal and connective tissue disorders (14 diseases), nervous system and developmental disorders (7 diseases), blood and immune system disorders (6 diseases), and cardiovascular and circulatory system disorders (4 diseases) being the most prevalent categories.

Among these diseases, we noticed that some are associated with cfDNA characteristics, including systemic lupus erythematosus (SLE, *DNASE1L3*), Barrett’s esophagus (BE, *MSR1*), tetralogy of Fallot (TOF, *ZFPM2*), autism (*RPL10*), congenital nongoitrous hypothyroidism (CHNG, *THRA*), amyotrophic lateral sclerosis (ALS, *TIA1*), and neurodevelopmental disorder with dysmorphic facies and distal skeletal anomalies (NEDDFSA, *ZMIZ1*). In detail, SLE is an autoimmune disease characterized by inflammation with the production of autoantibodies. There is increasing evidence that failure in the clearance of cfDNA by deoxyribonucleases (DNASES), particularly *DNASE1L3*, can lead to the generation of anti-DNA antibodies and SLE [72–74]. The plasma cfDNA levels are closely related to the disease severity of SLE and can potentially be used as a biomarker for SLE diagnosis. The gene *MSR1*, macrophage scavenger receptor 1, plays an essential inflammatory role in multiple processes, including immunity, lung and liver disease, and cancer [75]. Previous studies have reported that mutations in *MSR1* are associated with the presence of BE [76]. The cfDNA analysis becomes a promising approach for monitoring the neoplastic progression of BE [77]. The TOF is a congenital heart disease and the cfDNA technology can be used for disease screening by NIPT [78]. The autism and ALS are associated with inflammatory processes related to immune system dysfunctions and the cfDNA is becoming a potential diagnostic tool for these diseases [79–81].

The identification of these associations between motif-associated genes and genetic disorders suggests a complex interplay between cfDNA end motifs, genetic variations, and human health.

## Methods

### Experimental model and study participant details

#### Subjects

The participants were recruited from Wuhan Children’s Hospital during their routine pregnancy examinations from 2017 to 2020. A variety of tests were conducted throughout the entire duration of pregnancy, including non-invasive prenatal testing (NIPT), biochemistry assessments, oral glucose tolerance tests, ultrasound screenings, and more. Each pregnant participant provided informed consent before enrollment. This study received approval from the Institutional Review Boards (IRB) of both Wuhan Children’s Hospital (2021R062-E03) and the Bioethics and Biosafety of BGI (BGI-IRB 21088-T2). Additionally, authorization was obtained from the National Human Genetic Resources Management Office (Approval No. [2021] CJ2002). Our sole inclusion criterion was the availability of NIPT genotype data. In total, we included 38,668 pregnant women after quality control based on sequencing depth and mapping rate. These individuals were then used for genotype imputation.

#### Pregnancy phenotypes

Throughout the approximately 40-week gestation period, pregnant women undergo a range of examinations including blood routine, liver function, kidney function, and more. In our prior investigation, we curated 104 biochemical examination metrics and birth outcome indicators, conducting a comprehensive genome-wide association analysis [34]. A detailed list for providing the information of these phenotypes, including full name, abbreviation, and phenotypic category is provided in Table S7.

#### Independent cohort for replication study

To verify our GWAS findings of cfDNA molecular features, we performed a replication study in an independent cohort with 442 healthy participants. In detail, these participants were recruited in an outpatient department during their health examinations from 2021 to 2022 in the city of Shenzhen. Each participant provided informed consent before enrollment. This study received ethnical approval from the Institutional Review Boards of the Bioethics and Biosafety of BGI (BGI-IRB 21157-T2). The plasma cfDNA was used for whole genome sequencing, using DNBSEQ platform with paired-end 100 bp mode and an average sequencing depth of ∼35X.

#### Murine models

The animal study was approved by the Institutional Review Boards of Bioethics and Biosafety of BGI (BGI-IRB A24009) and the Institutional Animal Care and Use Committee (IACUC) of Cyagen (GACU24-SY028). The *Panx1*-deficient mice model (C57BL/6JCya) and *Dnase1l1*-deficient mice model (C57BL/6JCya) were created by CRISPR/Cas-mediated genome engineering at Cyagen. The knockout of genes was confirmed by PCR and sequencing. Housing conditions include a 12-hour light-dark cycle, controlled temperature and humidity, and pelleted rodent chow. 8-week-old male and female mice were used for the experiment. Gender differences were not analyzed for *Panx1*-deficient mice. *Dnase1l1* exists on the X chromosome. *Dnase1l1*-deficient mice were compared among groups of female homozygous, male hemizygous, and female heterozygous.

#### Cell line models

*PANX1* knockout Jurkat cell line model and *DNASE1L1* knockout HEK293 cell line model were created by CRISPR/Cas-mediated genome engineering at Cyagen.

### Method details

#### Whole-genome sequencing

The DNA samples used for sequencing were obtained from peripheral blood collected from pregnant women for NIPT testing, which contains both maternal and fetal cell-free DNA. Since NIPT testing is typically conducted around 16 weeks of gestation when the fetal DNA content is approximately 10%, with half of it being identical to maternal DNA, we did not differentiate between maternal and fetal DNA in subsequent analyses. Instead, we treated all DNA as maternal for analysis purposes. Using the BGISEQ 500 sequencing platform, we employed a combination of probe anchoring and polymerase sequencing for single-end sequencing, with reads length of 35 bp.

We employed the fastp software [82] to conduct quality control analysis on the sequencing data stored in FASTQ format. This involved the removal of adapter sequences and elimination of low-quality sequence fragments (reads). Following this, we utilized BWA [83] to align the quality-controlled reads to the hg38 reference genome [84], subsequently converting the aligned reads into BAM format and sorting them. Duplicate reads were subsequently eliminated from the sorted BAM files using the Samtools rmdup tool [85]. Additionally, we utilized the GATK [86] BaseRecalibrator to recalibrate base quality scores (BQSR) on the sorted BAM files using known site information. The GATK ApplyBQSR tool was then utilized to perform further base quality score recalibration and sorting on the sorted BAM files, thereby generating index files. Finally, Samtools stats was employed to generate comprehensive statistics for the calibrated BAM files.

#### Variant detection and Genotype imputation

Given the ultra-low nature of NIPT sequencing data, traditional analysis workflows designed for high-depth sequencing, such as variant detection, are inadequate. To ensure precise variant discovery, we used BaseVar [87], which was developed for calling variants in ultra-low-pass (<1.0x) sequencing data. Additionally, for inferring missing genotypes, we used the STITCH software [88] with entering the sequence bam files as input.

#### CfDNA end motifs

In accordance with the previous definition. Four mer ‘end motif’ in this study refers to the initial 4 nucleotides at the 5’ end of each strand of plasma DNA molecules [16]. The occurrence of each of the 256 motifs was calculated to determine motif frequency.

#### Murine sample collection

Mice were killed by removing eyeballs and blood was collected in EDTA tubes (KANGJIAN). In the *Panx1* module, 6 *Panx1*−/− mice (3 males and 3 females) and 6 WT mice (3 males and 3 females) were used. In the *Dnase1l1* module, 5 homozygous female, 5 heterozygous female, and 5 hemizygous male mice were used. The blood was first centrifuged at 1600 g for 10 min at 4 ◦C to collect the upper layer of plasma, and then the plasma was centrifuged at 16,000 g for 10 min at 4 ◦C to remove cellular debris. The supernatant plasma samples were stored at –80 ◦C before cfDNA extraction.

#### Cell line culture

Gene knockout cell lines were confirmed by sequencing. WT and *PANX1* knockout cells were cultured in RPMI 1640 medium, and *DNASE1L1* knockout cells were cultured in DEME medium at 37 ◦C in a humidified atmosphere with 5% CO2. 10% fetal bovine serum was added to the medium. The cell culture supernatant was first centrifuged at 1,600 g for 10 min at 4 ◦C, and then centrifuged again at 16,000 g for 10 min at 4 ◦C to remove cells and cellular debris during sample preparation.

#### Cell-free DNA extraction and quantification

The blood plasma and cell culture supernatant samples were used for cell-free DNA extraction using MagPure Circulating DNA KF Kit (MD5432-02, Magen). 100 μL plasma from each mouse and 500 μL cell culture supernatant for each sample were used for cfDNA extraction. The cfDNA concentrations were measured by Qubit™ 4.0 Fluorometer (Invitrogen) using Qubit dsDNA High Sensitivity Assay Kit (Q32854, Invitrogen).

### Quantification and statistical analysis

#### Genome-wide association study

We utilized the genotype dosage (a quantitative value between 0 and 2) imputed by STITCH as genotype data, and 256 cfDNA end motifs served as phenotypes for the genome-wide association analysis. To control for population stratification, we performed principal component analysis (PCA) on the population genetic variation identified by BaseVar using PLINK2 [89], selecting the top 10 principal components as covariates. Furthermore, maternal age, gestational age at NIPT testing, and sequencing depth were included as additional covariates. The GWAS analysis was performed by using PLINK2, which is capable of handling dosage data. We filter in SNPs with minor allele frequency (MAF) greater than 0.05, p-value for testing Hardy-Weinberg equilibrium (HWE) greater than 1e-6, and genotype missingness less than 10%. We set the significance threshold at 5e-8 for genome-wide significance and 1.95e-10 (=5e-8/256) for study-wide significance. In the independent replication study, we used PLINK2 to perform the GWAS analysis by adjusting age, gender, and the first 10 PCs of genotype as covariates. The SNPs with MAF > 0.01 and HWE p-value > 1e-6 were filtered in the GWAS analysis.

#### Heritability partition

To further understand the genetic architecture of the cfDNA molecular features and the polygenic contributions to heritability of different genetic components, including cell type–specific elements, we performed a heritability partition analysis. Specifically, to partition heritability from GWAS summary statistics of the 256 end motifs, we used stratified LD score regression (stratified LDSC) [90], which can account for linkage disequilibrium (LD). The cell type-specific expression data are referred to the Genotype-Tissue Expression (GTEx) project [91] and Franke lab dataset [92]. In total, there are 205 tissues and cell types from nine categories, including adipose, blood/immune, cardiovascular, central nervous system, digestive, endocrine, liver, musculoskeletal/connective, and others. The LD reference data is the 1000 Genomes Phase 3 East Asian populations.

#### Pathway-based analysis

Pathway enrichment analysis assists in highlighting pathways enriched with candidate genes pivotal for biological functions, thereby facilitating in unraveling mechanistic insights into the underlying traits. Leveraging summary statistics from GWAS conducted on 256 motifs, we performed pathway enrichment analysis using PASCAL (Pathway scoring algorithm) software—an intuitive tool designed for gene scoring and pathway analysis from GWAS results [93]. The databases include Reactome [94], Biocarta database (http://www.biocarta.com/), and KEGG [95]. For each of the 256 motifs, we computed the median pathway score and prioritized pathways in ascending order based on their scores, with special attention to those with higher rankings.

#### Genetic correlation

Genetic correlation is defined as an informative metric to quantify the overall genetic similarity between two complex traits. In this section, we investigated the genetic correlation between 256 end motifs and 104 pregnancy phenotypes measured in the same group of individuals. Specifically, we used LD score (LDSC) regression to perform the genetic correlation analysis with reference panel being East Asian population from the 1000 Genomes Project [96].

#### Mendelian randomization

Mendelian randomization (MR) is a methodological approach used in epidemiology and genetics to investigate causal relationships between exposure factors and outcomes. In this study, we propose performing bi-directional MR analysis using end motifs and pregnancy phenotypes as either exposure or outcome variables. Specifically, we conducted LD clumping to select only independent SNPs within a 10,000 kb window and a clumping r^2^ threshold of 0.1. LD was calculated based on the East Asian population from the 1000 Genomes Project. To infer causality, we employed the inverse variance weighted (IVW) method [97] and MR-Egger method [98] using the R function *TwoSampleMR*:mr [99]. It is worth noting that we only selected exposure variables with more than three significant SNPs after LD clumping to ensure the reliability of the MR analysis. We used a suggestive significance threshold of 1e-3 based on both IVW method and MR-Egger method.

#### Colocalization analysis

Colocalization analysis is a statistical method used to determine whether two or more traits share the same causal genetic variant(s) within a specific genomic region. This approach is essential when different GWAS identify overlapping signals, suggesting a potential shared genetic architecture between traits. To investigate whether the cfDNA end motifs and pregnancy phenotypes have overlapped genetic effects due to the same causal variant, we performed colocalization analysis using R package *coloc* [100]. The prior probability of a SNP is associated with either end motif or phenotype is set to be 1e-4 and with both traits is 1e-5. For visualizations of the colocalized regions between end motifs and phenotypes, we used R package *locuscomparer* [101].

#### Pleiotropy analysis with public databases

In this section, we investigated the pleiotropic effects of motif-associated genes based on GWAS catalog [31] and OMIM compendium [102]. First, we download SNP-trait association data from the GWAS catalog and proceed to process and analyze them through a series of steps, ultimately visualizing the distribution of traits. Initially, we download the latest association data from the GWAS catalog and perform initial filtering, retaining only statistically significant SNPs at the whole-genome level, with associations having a p-value less than 5e-8. Following this, we define a search region based on predefined loci associated with cfDNA motifs, extending 100 kb upstream and downstream. For each identified search region, we identify all significant SNPs and remove duplicates based on the PubMed ID and associated trait to ensure each trait is recorded only once per study. Then, for each trait, we count the total number of their associated loci in GWAS catalog and the number of loci shared with end motifs, and calculated the ratio. Finally, we visualize the distribution of traits using bubble plots.

OMIM (Online Mendelian Inheritance in Man) is a comprehensive database of human genes and genetic disorders. We created a list for all mapped genes of SNPs associated with cfDNA end motifs and searched their related diseases in the OMIM database.

## Discussion

cfDNA molecular features, such as fragment size, end motifs, and methylation patterns, are vital for understanding various biological processes and disease states. These characteristics of cfDNA molecules have also revolutionized liquid biopsy technology, allowing for non-invasive diagnosis, monitoring disease progression, and predicting patient outcomes. Uncovering novel key genes involved in cfDNA generation and shaping of cfDNA characteristics offers valuable insights into cfDNA biology and potential therapeutic targets for cfDNA-related diseases. However, there are only a very limited number of studies investigating the genetic mechanisms underlying cfDNA molecular signatures.

In this study, we retrieved the sequencing data from the NIPT of 28,016 Chinese pregnant women. We utilized whole-genome sequencing data of cfDNA to extract both genotype information and cell-free DNA features. For the first time, we conducted a genome-wide association study to explore the genetic basis of cfDNA molecular features. In total, we identified 84 genetic loci associated with 218 motifs. The most significant loci include *PANX1*, *DFFB*, *DNASE1L3*, *DNASE1L1*, and *PSMD3*. These findings were further validated through independent GWAS analyses and experimental designs, strengthening their biological significance. By in-depth comparison of the cfGWAS results with our parallel pregnancy phenotype GWAS study and the GWAS catalog, we observed genetic correlations between BMI, weight, uric acid, and cfDNA. Additionally, we revealed a strong causal relationship between hematological factors, especially leukocytes, and cfDNA features.

Although the sample size in our study does not reach the scale of hundreds of thousands, cfDNA screenings such as NIPT, tumor liquid biopsies, and pathogen liquid biopsies have generated vast amounts of genomic data. Consequently, the potential of our cfGWAS paradigm is enormous. For example, by June 2023, over 40 million pregnant women worldwide had undergone NIPT testing, resulting in a substantial amount of sequence data. This sample volume far exceeds that of participants with general WGS data [103]. In addition, understanding the biology of cfDNA generation and clearance could highlight potential applications in the field of liquid biopsy. For instance, circulating tumor DNA (ctDNA) is often present in low quantities, making it challenging to collect sufficient blood for accurate detection, particularly in cases with small tumors. Therefore, strategies to protect circulating DNA from degradation or to reduce its clearance could ensure adequate concentrations of ctDNA, enabling sensitive and robust liquid biopsy tests even with small blood volumes [104]. Furthermore, our research could contribute to the discovery of therapeutic targets for diseases influenced by abnormal DNA concentrations, such as SLE and gout. This includes insights into drug targets that accelerate the clearance of circulating DNA.

Given the widespread clinical use of cfDNA, the significant discoveries from cfGWAS and their potential applications will not only revolutionize our understanding of cfDNA biology but also open new avenues for future clinical applications.

## Limitations of study

Limited by the sequencing strategy of NIPT technology (single-end 35bp), we only used end motifs as a snapshot for cfDNA features. Although different cfDNA features were strongly correlated, a cfGWAS of other features, including fragment length, concentration, jagged ends, and nucleosome footprints, would provide a more comprehensive investigation. Furthermore, our discovery set relied on a cohort of pregnant women, so it’s unclear how many findings are general for all people or specific to pregnancy. Independent studies on non-pregnant cohorts are necessary to resolve this uncertainty. In addition, our discovery GWAS study had a relatively small sample size of 20,000 pregnant women. The statistical power of GWAS is heavily dependent on sample size, with larger samples yielding higher powers to detect genetic signals. We acknowledge the importance of amassing larger sample sizes for more robust and comprehensive genetic investigations in the future. We anticipate uncovering even more novel genetic associations that can further illuminate the complex genetic basis of cfDNA features. In this study, we only validated the impact of *PANX1* and *DNASE1L1* genes on cfDNA concentration using knockout mouse models and cell lines. Further studies are needed to fully uncover the biological mechanisms of these genes on cfDNA. Validation of other candidate genes in knockout models would also be necessary.

## Data Availability

The data that support the findings of this study have been deposited into CNGB Sequence Archive (CNSA) of China National GeneBank DataBase (CNGBdb) with accession number CNP0005734. Any additional information required to reanalyze the data reported in this paper is available from the lead contact upon request.

## Acknowledgment

This study was supported by National Natural Science Foundation of China (No.32171441), National Key Research and Development Program of China (No.2023YFC2605400, No.2022YFC2502402), Guangzhou Basic and Applied Basic Research Foundation (No.202201010189), Open Project of State Key Laboratory of Respiratory Disease (No.SKLRD-OP-202309), the Innovation Platform for Academicians of Hainan Province (No.YSPTZX202118), Key-Area Research and Development Program of Guangdong Province (No.2023B0303040001), the Shenzhen Key Laboratory of Genomics (No.CXB200903110066A), Guangdong Provincial Key Laboratory of Human Disease Genomics (No.2020B1212070028), Guangdong Provincial Key Laboratory of Genome Read and Write (No.2017B030301011), and the China National GeneBank.

## Author contribution

X.J., H.Q.Z. and A.Z. conceived the study, designed the research program, and managed the project.

H.X., M.Y., and A.Z. collected the data.

L.L., J.Z. and Y.W. preprocessed the data and finished the quality control.

H.H.Z., L.L., R.O., X.Z., Yu L., Ying L., L.W., G.Z., L.G.L. performed the statistical analysis and results visualization.

H.Q.Z., Y.Z., S.Z., F.L., and C.X. analyzed the experimental data.

X.J., H.H.Z., H.Q.Z, Y.Z., S.Z., L.L., R.O., X.Z. and Ying L. wrote the manuscript.

All authors participated in revising the manuscript.

## Resource availability

### Data and code availability

The data that support the findings of this study have been deposited into CNGB Sequence Archive (CNSA) [105] of China National GeneBank DataBase (CNGBdb) [106] with accession number CNP0005734. Any additional information required to reanalyze the data reported in this paper is available from the lead contact upon request.

## Figures and tables

**Figure S1.** The QQ-plots and genomic inflation factors of GWAS results for 256 motifs

**Figure S2.** The regional plot of the top 4 GWAS signals

**Figure S3.** The Manhattan plot and regional plots of the independent validation dataset

**Figure S4.** The heritability and number of significant loci of 256 motifs

**Figure S5.** The results for partitional heritability analysis

**Figure S6.** The significant count and p-value distribution of pathway-based analysis

**Figure S7.** The count of significantly genetic correlated motifs of each phenotype

**Figure S8.** The MR regression lines of the 14 casually related phenotype

**Figure S9.** Colocalization regional plot for 6 loci

**Table S1.** Aggregated GWAS results of motif-associated loci

**Table S2.** The heritability results for each motif

**Table S3.** The p-value for partitioned heritability analysis

**Table S4.** The significant count for partitioned heritability analysis

**Table S5.** The p-value for pathway-based analysis

**Table S6.** The significant count for pathway-based analysis

**Table S7.** The genetic correlation rg-value between motifs and pregnancy phenotypes

**Table S8.** The genetic correlation p-value between motifs and pregnancy phenotypes

**Table S9.** The significant count for genetic correlation

**Table S10.** The results for Mendelian Randomization analysis

**Table S11.** The results of sensitivity tests for causal effects in Mendelian randomization

**Table S12.** The colocalization results between motifs and phenotypes

**Table S13.** The results for pleiotropy analyses with associations from GWAS catalog

**Table S14.** The genes associated with both motifs and diseases from OMIM database

## References

1. Wan, J.C.M., et al., Liquid biopsies come of age: towards implementation of circulating tumour DNA. Nat Rev Cancer, 2017. 17(4): p. 223–238.

2. Chiu, R.W., et al., Non-invasive prenatal assessment of trisomy 21 by multiplexed maternal plasma DNA sequencing: large scale validity study. Bmj, 2011. 342: p. c7401.

3. Cisneros-Villanueva, M., et al., Cell-free DNA analysis in current cancer clinical trials: a review. Br J Cancer, 2022. 126(3): p. 391–400.

4. Medina, J.E., et al., Cell-free DNA approaches for cancer early detection and interception. J Immunother Cancer, 2023. 11(9).

5. Knight, S.R., A. Thorne, and M.L. Lo Faro, Donor-specific Cell-free DNA as a Biomarker in Solid Organ Transplantation. A Systematic Review. Transplantation, 2019. 103(2): p. 273–283.

6. Chan, R.W., et al., Plasma DNA aberrations in systemic lupus erythematosus revealed by genomic and methylomic sequencing. Proc Natl Acad Sci U S A, 2014. 111(49): p. E5302–11.

7. Nomura, J., et al., Rapid detection of invasive Mycobacterium chimaera disease via a novel plasma-based next-generation sequencing test. BMC Infect Dis, 2019. 19(1): p. 371.

8. Zheng, Y., et al., Development and clinical validation of a droplet digital PCR assay for detecting Acinetobacter baumannii and Klebsiella pneumoniae in patients with suspected bloodstream infections. Microbiologyopen, 2021. 10(6): p. e1247.

9. Qi, T., et al., Cell-Free DNA Fragmentomics: The Novel Promising Biomarker. Int J Mol Sci, 2023. 24(2).

10. Zhou, Z., et al., Fragmentation landscape of cell-free DNA revealed by deconvolutional analysis of end motifs. Proc Natl Acad Sci U S A, 2023. 120(17): p. e2220982120.

11. Koukourakis, M.I., et al., Circulating Plasma Cell-free DNA (cfDNA) as a Predictive Biomarker for Radiotherapy: Results from a Prospective Trial in Head and Neck Cancer. Cancer Diagn Progn, 2023. 3(5): p. 551–557.

12. Li, L., et al., Serum Cell-Free DNA-based Detection of Mycobacterium avium Complex Infection. Am J Respir Crit Care Med, 2024. 209(10): p. 1246–1254.

13. Lehmann, J., et al., Plasma mtDNA as a possible contributor to and biomarker of inflammation in rheumatoid arthritis. Arthritis Res Ther, 2024. 26(1): p. 97.

14. Lapin, M., et al., Fragment size and level of cell-free DNA provide prognostic information in patients with advanced pancreatic cancer. J Transl Med, 2018. 16(1): p. 300.

15. Thierry, A.R., Circulating DNA fragmentomics and cancer screening. Cell Genom, 2023. 3(1): p. 100242.

16. Jiang, P., et al., Plasma DNA End-Motif Profiling as a Fragmentomic Marker in Cancer, Pregnancy, and Transplantation. Cancer Discov, 2020. 10(5): p. 664–673.

17. Moser, T., et al., Bridging biological cfDNA features and machine learning approaches. Trends Genet, 2023. 39(4): p. 285–307.

18. Zhu, G., et al., Tissue-specific cell-free DNA degradation quantifies circulating tumor DNA burden. Nat Commun, 2021. 12(1): p. 2229.

19. Jacobson, M.D., M. Weil, and M.C. Raff, Programmed cell death in animal development. Cell, 1997. 88(3): p. 347–54.

20. Mannherz, H.G., et al., A new function for an old enzyme: the role of DNase I in apoptosis. Curr Top Microbiol Immunol, 1995. 198: p. 161–74.

21. Rodriguez, A.M., et al., Identification, localization, and expression of two novel human genes similar to deoxyribonuclease I. Genomics, 1997. 42(3): p. 507–13.

22. Widlak, P., et al., Cleavage preferences of the apoptotic endonuclease DFF40 (caspase-activated DNase or nuclease) on naked DNA and chromatin substrates. J Biol Chem, 2000. 275(11): p. 8226–32.

23. Han, D.S.C. and Y.M.D. Lo, The Nexus of cfDNA and Nuclease Biology. Trends Genet, 2021. 37(8): p. 758–770.

24. Sin, S.T., et al., Effects of nucleases on cell-free extrachromosomal circular DNA. JCI Insight, 2022. 7(8).

25. Chen, M., et al., Fragmentomics of urinary cell-free DNA in nuclease knockout mouse models. PLoS Genet, 2022. 18(7): p. e1010262.

26. Han, D.S.C., et al., The Biology of Cell-free DNA Fragmentation and the Roles of DNASE1, DNASE1L3, and DFFB. Am J Hum Genet, 2020. 106(2): p. 202–214.

27. Uffelmann, E., et al., Genome-wide association studies. Nature Reviews Methods Primers, 2021. 1(1): p. 59.

28. Zhang, H., et al., Genome-wide association study identifies 32 novel breast cancer susceptibility loci from overall and subtype-specific analyses. Nat Genet, 2020. 52(6): p. 572–581.

29. Tcheandjieu, C., et al., Large-scale genome-wide association study of coronary artery disease in genetically diverse populations. Nat Med, 2022. 28(8): p. 1679–1692.

30. Borrego-Yaniz, G., et al., Risk loci involved in giant cell arteritis susceptibility: a genome-wide association study. Lancet Rheumatol, 2024. 6(6): p. e374–e383.

31. Sollis, E., et al., The NHGRI-EBI GWAS Catalog: knowledgebase and deposition resource. Nucleic Acids Res, 2023. 51(D1): p. D977–d985.

32. Liu, S., et al., Genomic Analyses from Non-invasive Prenatal Testing Reveal Genetic Associations, Patterns of Viral Infections, and Chinese Population History. Cell, 2018. 175(2): p. 347–359.e14.

33. Liu, S., et al., Utilizing Non-Invasive Prenatal Test Sequencing Data Resource for Human Genetic Investigation. bioRxiv, 2023: p. 2023.12.11.570976.

34. Xiao, H., et al., Genetic analysis of 104 pregnancy phenotypes in 39,194 Chinese women. medRxiv, 2023: p. 2023.11.23.23298979.

35. Huang, Q., et al., Association between genetic predisposition and disease burden of stroke in China: a genetic epidemiological study. Lancet Reg Health West Pac, 2023. 36: p. 100779.

36. Li, Z., et al., CMDB: the comprehensive population genome variation database of China. Nucleic Acids Res, 2023. 51(D1): p. D890–d895.

37. Whyte-Fagundes, P. and G. Zoidl, Mechanisms of pannexin1 channel gating and regulation. Biochim Biophys Acta Biomembr, 2018. 1860(1): p. 65–71.

38. Santavanond, J.P., et al., The small molecule raptinal can simultaneously induce apoptosis and inhibit PANX1 activity. Cell Death Dis, 2024. 15(2): p. 123.

39. Ueki, M., et al., Evaluation of all nonsynonymous single-nucleotide polymorphisms in the gene encoding human deoxyribonuclease I-like 1, possibly implicated in the blocking of endocytosis-mediated foreign gene transfer. DNA Cell Biol, 2014. 33(2): p. 79–87.

40. Blander, J.M., The many ways tissue phagocytes respond to dying cells. Immunol Rev, 2017. 277(1): p. 158–173.

41. MSigDB, G., Human Gene Set: BIOCARTA_SET_PATHWAY.

42. MSigDB, G., Human Gene Set: BIOCARTA_DNAFRAGMENT_PATHWAY.

43. Juul, L.A., et al., Noninvasive prenatal testing and maternal obesity: A review. Acta Obstet Gynecol Scand, 2020. 99(6): p. 744–750.

44. Mhatre, M., et al., The Effect of Maternal Obesity on Placental Cell-Free DNA Release in a Mouse Model. Reprod Sci, 2019. 26(9): p. 1218–1224.

45. Stupak, A., et al., The Influence of Maternal Obesity on Cell-Free Fetal DNA and Blood Pressure Regulation in Pregnancies with Hypertensive Disorders. Medicina (Kaunas), 2021. 57(9).

46. Wang, E., et al., Gestational age and maternal weight effects on fetal cell-free DNA in maternal plasma. Prenat Diagn, 2013. 33(7): p. 662–6.

47. Haghiac, M., et al., Increased death of adipose cells, a path to release cell-free DNA into systemic circulation of obese women. Obesity (Silver Spring), 2012. 20(11): p. 2213–9.

48. Lapaire, O., et al., Significant correlation between maternal body mass index at delivery and in the second trimester, and second trimester circulating total cell-free DNA levels. Reprod Sci, 2009. 16(3): p. 274–9.

49. Vora, N.L., et al., A multifactorial relationship exists between total circulating cell-free DNA levels and maternal BMI. Prenat Diagn, 2012. 32(9): p. 912–4.

50. Johnson, R.J., M.A. Lanaspa, and E.A. Gaucher, Uric acid: a danger signal from the RNA world that may have a role in the epidemic of obesity, metabolic syndrome, and cardiorenal disease: evolutionary considerations. Semin Nephrol, 2011. 31(5): p. 394–9.

51. Gherghina, M.E., et al., Uric Acid and Oxidative Stress-Relationship with Cardiovascular, Metabolic, and Renal Impairment. Int J Mol Sci, 2022. 23(6).

52. Li, D., et al., Reactive oxygen species induced by uric acid promote NRK–52E cell apoptosis through the NEK7–NLRP3 signaling pathway. Mol Med Rep, 2021. 24(4).

53. Liu, N., et al., Phosphatidylserine decarboxylase downregulation in uric acid–induced hepatic mitochondrial dysfunction and apoptosis. MedComm (2020), 2023. 4(4): p. e336.

54. Moss, J., et al., Comprehensive human cell-type methylation atlas reveals origins of circulating cell-free DNA in health and disease. Nat Commun, 2018. 9(1): p. 5068.

55. Leal, A., et al., White blood cell and cell-free DNA analyses for detection of residual disease in gastric cancer. Nat Commun, 2020. 11(1): p. 525.

56. Mattox, A.K., et al., The Origin of Highly Elevated Cell-Free DNA in Healthy Individuals and Patients with Pancreatic, Colorectal, Lung, or Ovarian Cancer. Cancer Discov, 2023. 13(10): p. 2166–2179.

57. Kustanovich, A., et al., Life and death of circulating cell-free DNA. Cancer Biol Ther, 2019. 20(8): p. 1057–1067.

58. Fridlich, O., et al., Elevated cfDNA after exercise is derived primarily from mature polymorphonuclear neutrophils, with a minor contribution of cardiomyocytes. Cell Rep Med, 2023. 4(6): p. 101074.

59. Moss, J., et al., Megakaryocyte– and erythroblast-specific cell-free DNA patterns in plasma and platelets reflect thrombopoiesis and erythropoiesis levels. Nat Commun, 2023. 14(1): p. 7542.

60. Medeiros, S.K., et al., Does cell-free DNA promote coagulation and inhibit fibrinolysis in patients with unprovoked venous thromboembolism? Thromb Res, 2020. 186: p. 13–19.

61. Sender, R., et al., What fraction of cellular DNA turnover becomes cfDNA? Elife, 2024. 12.

62. Liang, N., et al., Mature Red Blood Cells Contain Long DNA Fragments and Could Acquire DNA from Lung Cancer Tissue. Adv Sci (Weinh), 2023. 10(7): p. e2206361.

63. Fararjeh, A.S., et al., Proteasome 26S Subunit, non-ATPase 3 (PSMD3) Regulates Breast Cancer by Stabilizing HER2 from Degradation. Cancers (Basel), 2019. 11(4).

64. Kamatani, Y., et al., Genome-wide association study of hematological and biochemical traits in a Japanese population. Nat Genet, 2010. 42(3): p. 210–5.

65. Astle, W.J., et al., The Allelic Landscape of Human Blood Cell Trait Variation and Links to Common Complex Disease. Cell, 2016. 167(5): p. 1415–1429.e19.

66. Chen, M.H., et al., Trans-ethnic and Ancestry-Specific Blood-Cell Genetics in 746,667 Individuals from 5 Global Populations. Cell, 2020. 182(5): p. 1198–1213.e14.

67. Kachuri, L., et al., Genetic determinants of blood-cell traits influence susceptibility to childhood acute lymphoblastic leukemia. Am J Hum Genet, 2021. 108(10): p. 1823–1835.

68. Ferreira, M.A., et al., Shared genetic origin of asthma, hay fever and eczema elucidates allergic disease biology. Nat Genet, 2017. 49(12): p. 1752–1757.

69. Zhu, Z., et al., Shared genetics of asthma and mental health disorders: a large-scale genome-wide cross-trait analysis. Eur Respir J, 2019. 54(6).

70. Cao, J., et al., Lipid Metabolism Affects Fetal Fraction and Screen Failures in Non-invasive Prenatal Testing. Front Med (Lausanne), 2021. 8: p. 811385.

71. Kananen, L., et al., Circulating cell-free DNA in health and disease – the relationship to health behaviours, ageing phenotypes and metabolomics. Geroscience, 2023. 45(1): p. 85–103.

72. Al-Mayouf, S.M., et al., Loss-of-function variant in DNASE1L3 causes a familial form of systemic lupus erythematosus. Nat Genet, 2011. 43(12): p. 1186–8.

73. Gerovska, D. and M.J. Araúzo-Bravo, Systemic Lupus Erythematosus Patients with DNASE1L3·Deficiency Have a Distinctive and Specific Genic Circular DNA Profile in Plasma. Cells, 2023. 12(7).

74. Mathapathi, S. and C.Q. Chu, Contribution of Impaired DNASE1L3 Activity to Anti-DNA Autoantibody Production in Systemic Lupus Erythematosus. Rheumatol Immunol Res, 2022. 3(1): p. 17–22.

75. Gudgeon, J., J.L. Marín-Rubio, and M. Trost, The role of macrophage scavenger receptor 1 (MSR1) in inflammatory disorders and cancer. Front Immunol, 2022. 13: p. 1012002.

76. Orloff, M., et al., Germline mutations in MSR1, ASCC1, and CTHRC1 in patients with Barrett esophagus and esophageal adenocarcinoma. Jama, 2011. 306(4): p. 410–9.

77. Rumiato, E., et al., Detection of genetic alterations in cfDNA as a possible strategy to monitor the neoplastic progression of Barrett’s esophagus. Transl Res, 2017. 190: p. 16–24.e1.

78. Biró, O., J. Rigó, Jr., and B. Nagy, Noninvasive prenatal testing for congenital heart disease – cell-free nucleic acid and protein biomarkers in maternal blood. J Matern Fetal Neonatal Med, 2020. 33(6): p. 1044–1050.

79. Mendioroz, M., et al., Liquid biopsy: a new source of candidate biomarkers in amyotrophic lateral sclerosis. Ann Clin Transl Neurol, 2018. 5(6): p. 763–768.

80. Robichaud, P.P., et al., Circulating cell-free DNA as potential diagnostic tools for amyotrophic lateral sclerosis. Neurosci Lett, 2021. 750: p. 135813.

81. Shmarina, G.V., et al., Oxidized cell-free DNA as a stress-signaling factor activating the chronic inflammatory process in patients with autism spectrum disorders. J Neuroinflammation, 2020. 17(1): p. 212.

82. Chen, S., et al., *fastp: an ultra-fast all-in-one FASTQ preprocessor*. Bioinformatics, 2018. 34(17): p. i884–i890.

83. Li, H. and R. Durbin, Fast and accurate short read alignment with Burrows-Wheeler transform. Bioinformatics, 2009. 25(14): p. 1754–60.

84. Schneider, V.A., et al., Evaluation of GRCh38 and de novo haploid genome assemblies demonstrates the enduring quality of the reference assembly. Genome Res, 2017. 27(5): p. 849–864.

85. Li, H., et al., The Sequence Alignment/Map format and SAMtools. Bioinformatics, 2009. 25(16): p. 2078–9.

86. McKenna, A., et al., The Genome Analysis Toolkit: a MapReduce framework for analyzing next-generation DNA sequencing data. Genome Res, 2010. 20(9): p. 1297–303.

87. ShujiaHuang. basevar (GitHub). Available from: https://github.com/ShujiaHuang/basevar.

88. Davies, R.W., et al., Rapid genotype imputation from sequence without reference panels. Nat Genet, 2016. 48(8): p. 965–969.

89. Chang, C.C., et al., Second-generation PLINK: rising to the challenge of larger and richer datasets. GigaScience, 2015. 4(1).

90. Finucane, H.K., et al., Partitioning heritability by functional annotation using genome-wide association summary statistics. Nat Genet, 2015. 47(11): p. 1228–35.

91. *Human genomics*. The Genotype-Tissue Expression (GTEx) pilot analysis: multitissue gene regulation in humans. Science, 2015. 348(6235): p. 648–60.

92. Fehrmann, R.S., et al., Gene expression analysis identifies global gene dosage sensitivity in cancer. Nat Genet, 2015. 47(2): p. 115–25.

93. Alonso-Gonzalez, A., et al., Gene-based analysis of ADHD using PASCAL: a biological insight into the novel associated genes. BMC Med Genomics, 2019. 12(1): p. 143.

94. Milacic, M., et al., The Reactome Pathway Knowledgebase 2024. Nucleic Acids Res, 2024. 52(D1): p. D672–d678.

95. Kanehisa, M. and S. Goto, KEGG: kyoto encyclopedia of genes and genomes. Nucleic Acids Res, 2000. 28(1): p. 27–30.

96. Bulik-Sullivan, B.K., et al., LD Score regression distinguishes confounding from polygenicity in genome-wide association studies. Nat Genet, 2015. 47(3): p. 291–5.

97. Burgess, S., A. Butterworth, and S.G. Thompson, Mendelian randomization analysis with multiple genetic variants using summarized data. Genet Epidemiol, 2013. 37(7): p. 658–65.

98. Bowden, J., G. Davey Smith, and S. Burgess, Mendelian randomization with invalid instruments: effect estimation and bias detection through Egger regression. Int J Epidemiol, 2015. 44(2): p. 512–25.

99. Hemani, G., et al., The MR-Base platform supports systematic causal inference across the human phenome. Elife, 2018. 7.

100. Giambartolomei, C., et al., Bayesian test for colocalisation between pairs of genetic association studies using summary statistics. PLoS Genet, 2014. 10(5): p. e1004383.

101. Liu, B., et al., Abundant associations with gene expression complicate GWAS follow-up. Nat Genet, 2019. 51(5): p. 768–769.

102. Hamosh, A., et al., Online Mendelian Inheritance in Man (OMIM), a knowledgebase of human genes and genetic disorders. Nucleic Acids Res, 2005. 33(Database issue): p. D514–7.

103. Shendure, J., G.M. Findlay, and M.W. Snyder, Genomic Medicine-Progress, Pitfalls, and Promise. Cell, 2019. 177(1): p. 45–57.

104. Martin-Alonso, C., et al., Priming agents transiently reduce the clearance of cell-free DNA to improve liquid biopsies. Science, 2024. 383(6680): p. eadf2341.

105. Guo, X., et al., CNSA: a data repository for archiving omics data. Database (Oxford), 2020. 2020.

106. Chen, F.Z., et al., CNGBdb: China National GeneBank DataBase. Yi Chuan, 2020. 42(8): p. 799–809.

