## Supplementary figures for "cfGWAS reveal genetic basis of cell-free DNA features"

**Figure S1.** The QQ-plots and genomic inflation factors of GWAS results for 256 motifs

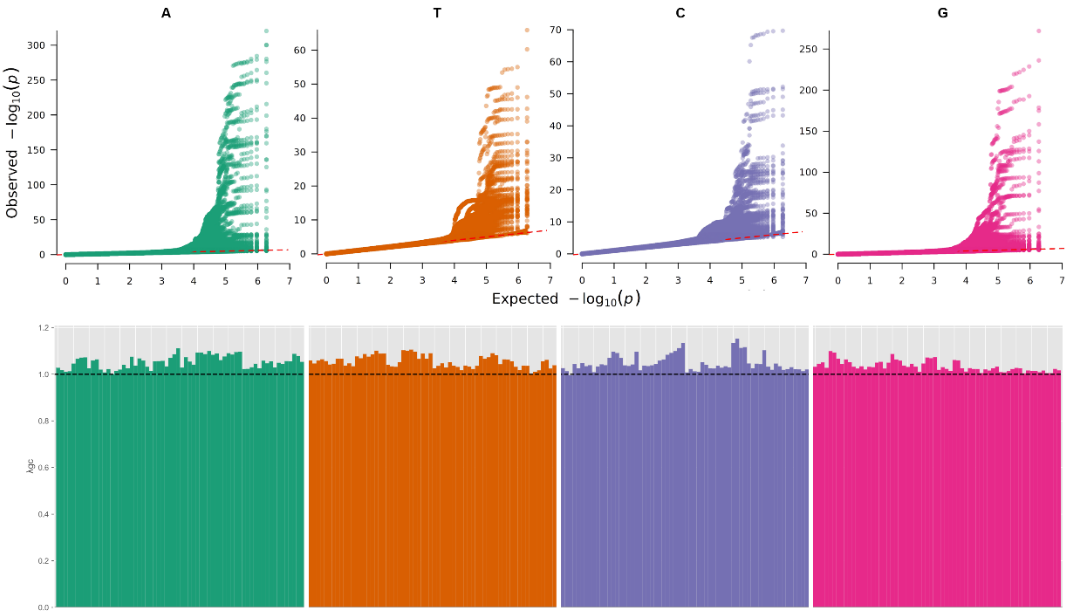

**Figure S2.** The regional plot of the top 4 GWAS signals

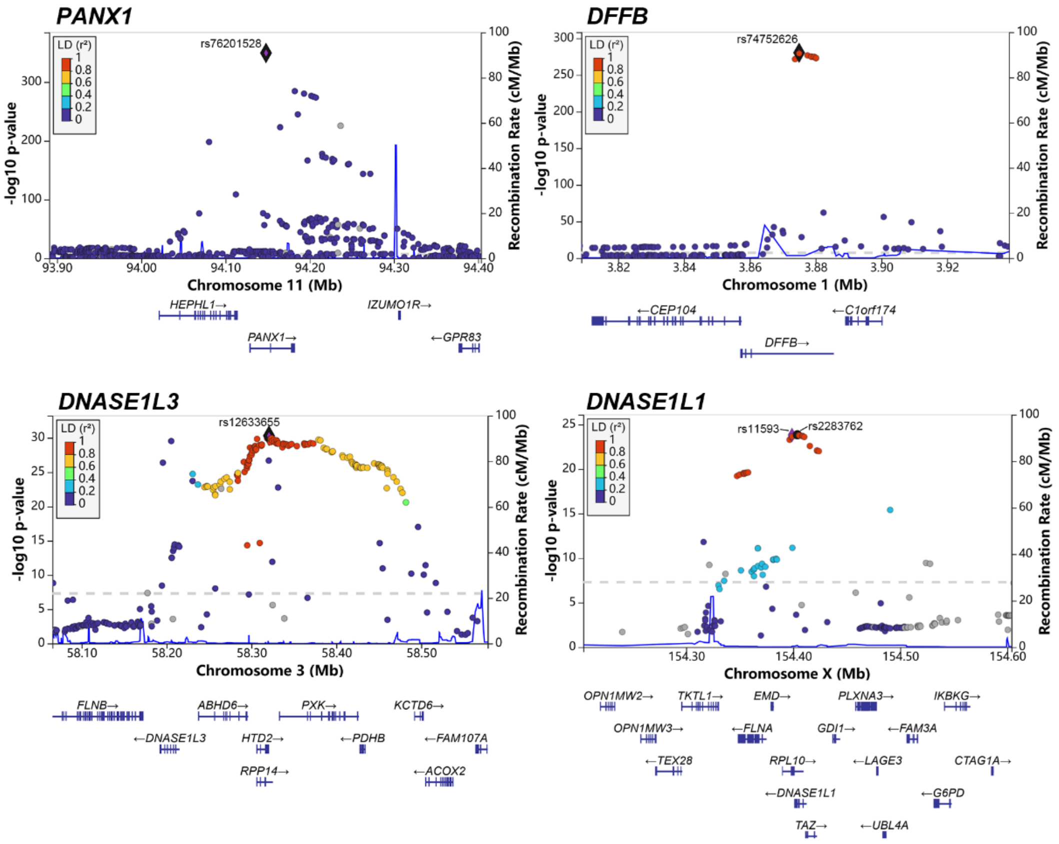

**Figure S3.** The Manhattan plot and regional plots of the independent validation dataset

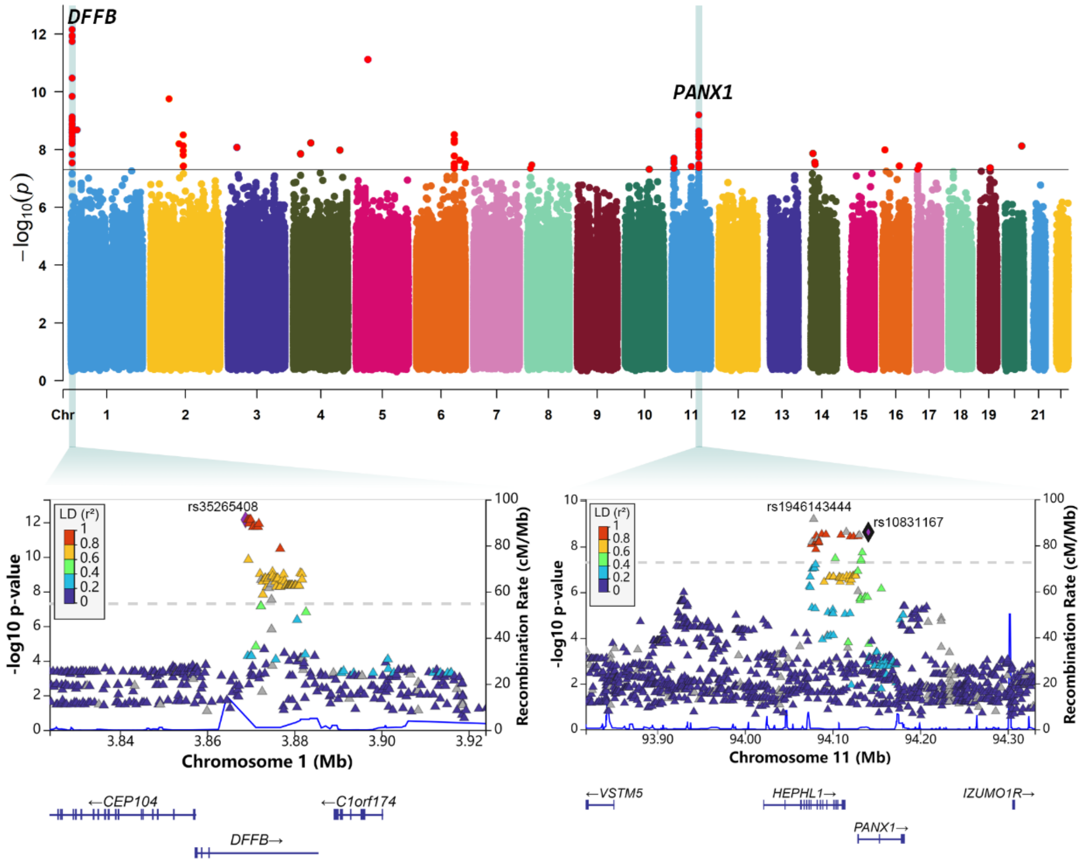

**Figure S4.** The heritability and number of significant loci of 256 motifs

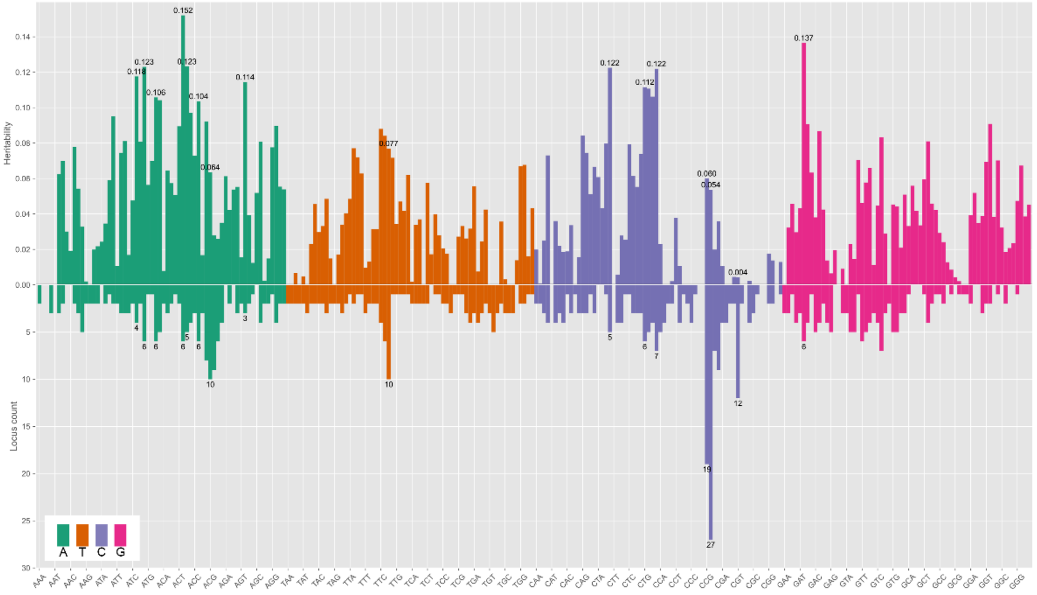

**Figure S5.** The results for partitional heritability analysis

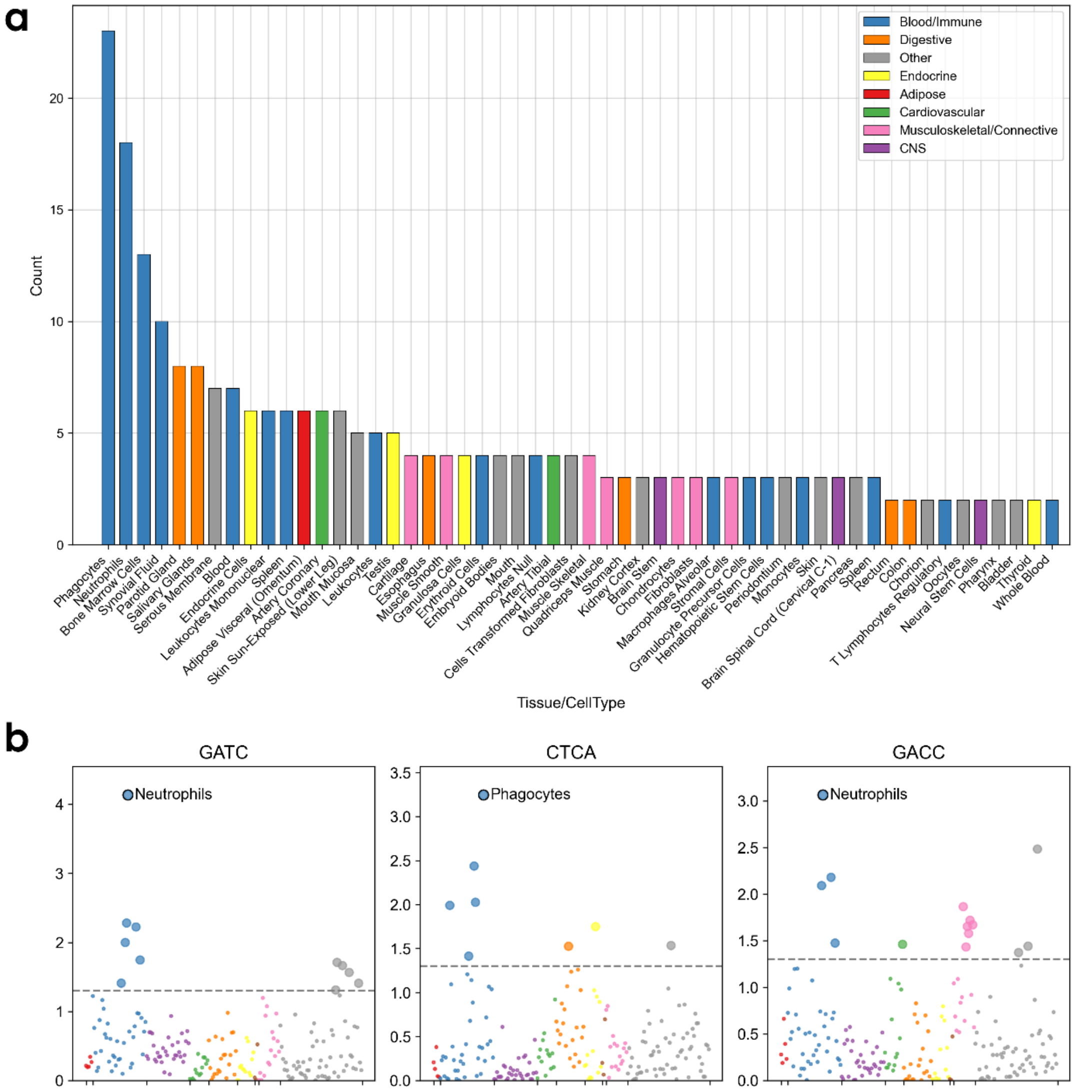

**Figure S6.** The significant count and p-value distribution of pathway-based analysis

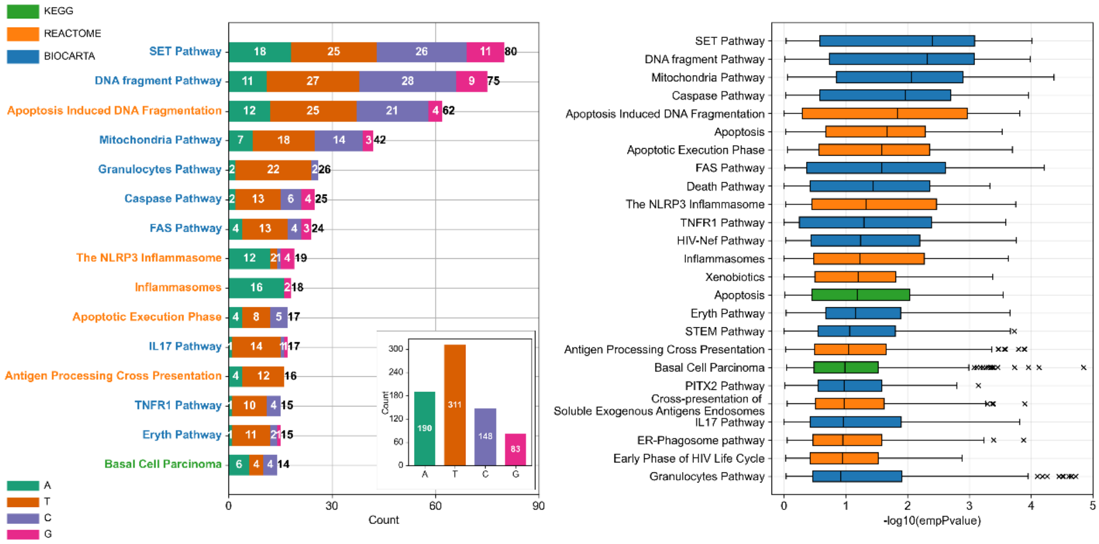

**Figure S7.** The count of significantly genetic correlated motifs of each phenotype

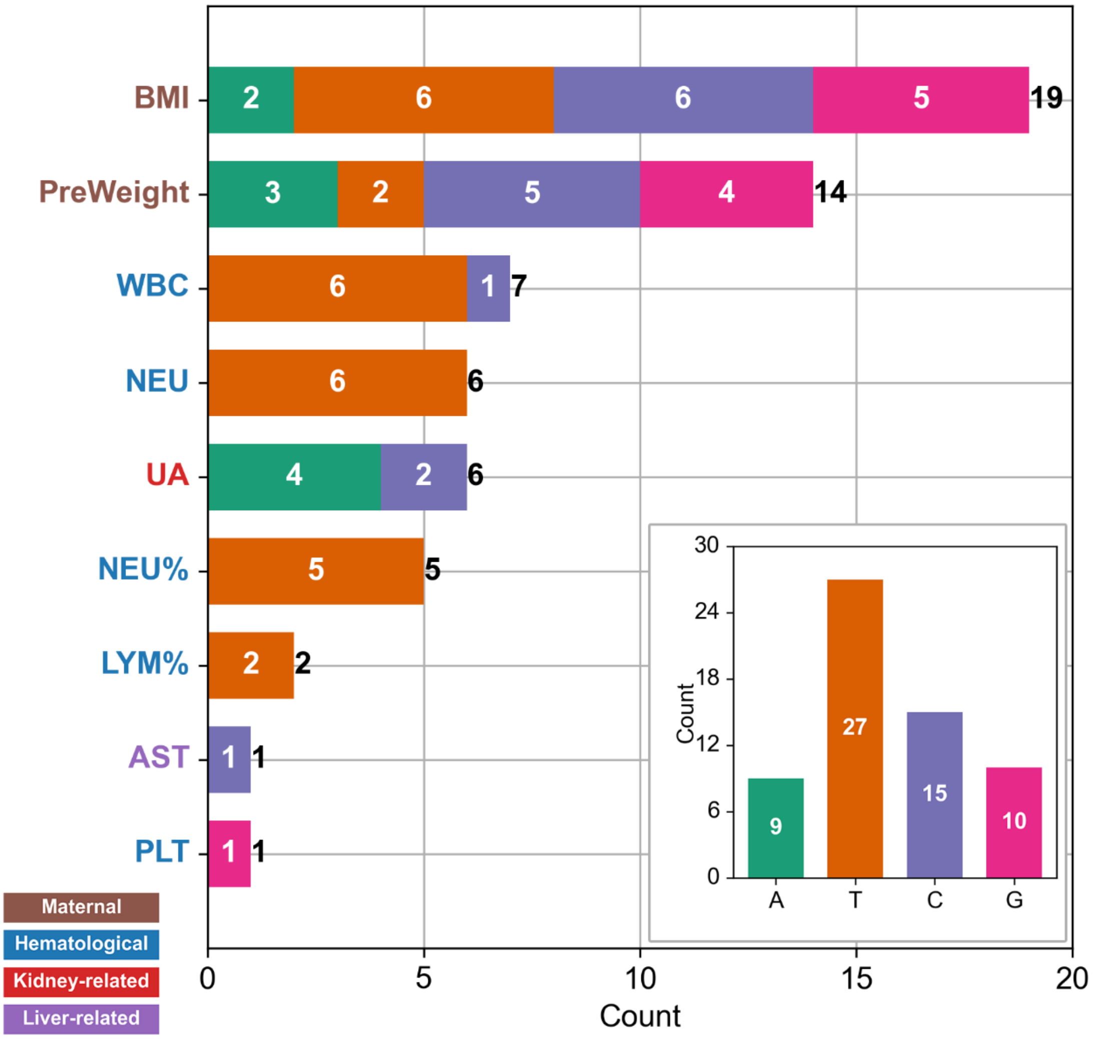

**Figure S8.** The MR regression lines of the 14 casually related phenotype

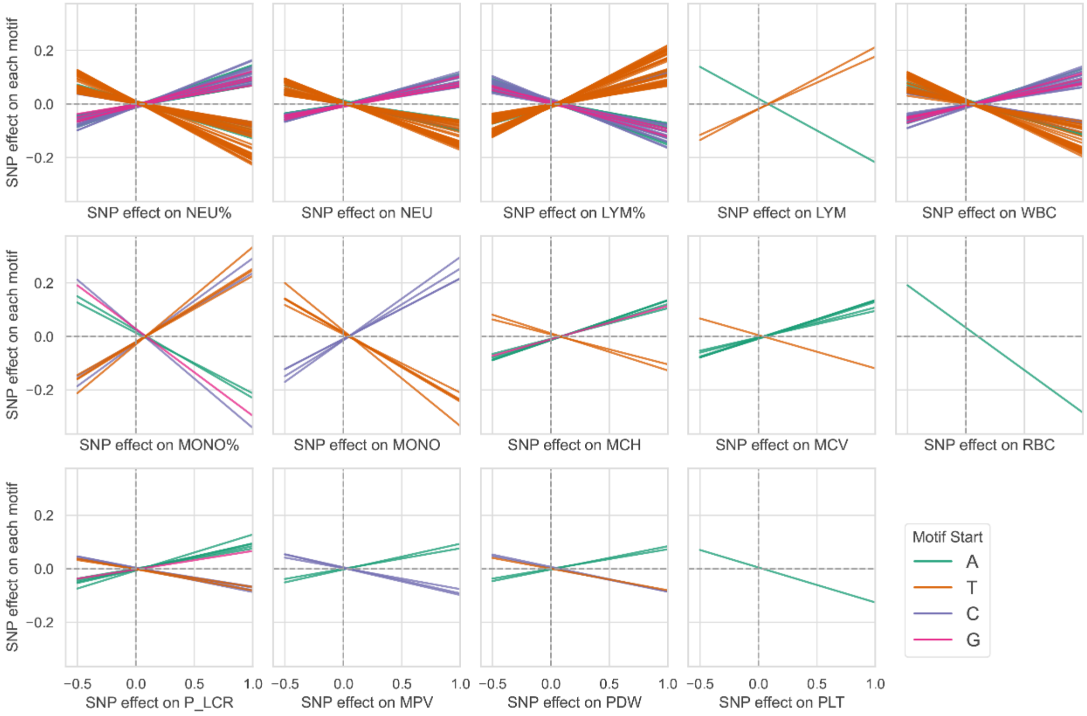

**Figure S9.** Colocalization regional plot for 6 loci

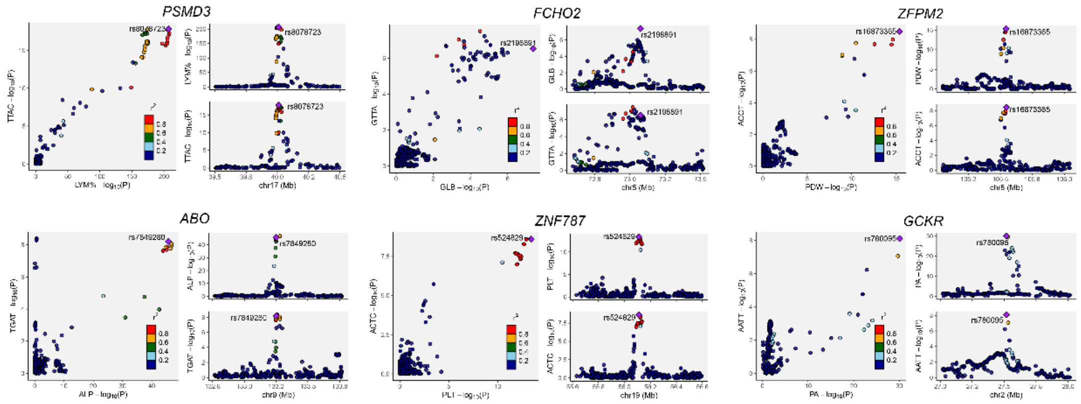
